# COVID-19 Vaccination Mandates and Vaccine Uptake

**DOI:** 10.1101/2021.10.21.21265355

**Authors:** Alexander Karaivanov, Dongwoo Kim, Shih En Lu, Hitoshi Shigeoka

## Abstract

We evaluate the impact of government-mandated proof of vaccination requirements for access to public venues and non-essential businesses on COVID-19 vaccine uptake. We find that the announcement of a mandate is associated with a rapid and significant surge in new vaccinations (more than 60% increase in weekly first doses), using the variation in the timing of these measures across Canadian provinces in a difference-in-differences approach. Time-series analysis for each province and for France, Italy and Germany corroborates this finding. Counterfactual simulations using our estimates suggest the following cumulative gains in the vaccination rate among the eligible population (age 12 and over) as of October 31, 2021: up to 5 percentage points (p.p.) (90% CI 3.9–5.8) for Canadian provinces, adding up to 979,000 (425,000-1,266,000) first doses in total for Canada (5 to 13 weeks after the provincial mandate announcements), 8 p.p. (4.3–11) for France (16 weeks post-announcement), 12 p.p. (5–15) for Italy (14 weeks post-announcement) and 4.7 p.p. (4.1–5.1) for Germany (11 weeks post-announcement).

## Introduction

Immunization has proven very effective for reducing the spread and severity of COVID-19, with large reductions in the risk of severe outcomes for vaccinated people, [1–4]. Yet, following a rapid uptake in early 2021, vaccinations in many countries (see Extended Data Fig. 1) slowed down significantly in the summer months. In addition, even locations with high vaccination rates experienced increased viral transmission or had to maintain or re-introduce non-pharmaceutical interventions such as mask wearing or indoor capacity limits in fall 2021 because of the elevated reproduction rate of the Delta variant.

Achieving high COVID-19 vaccination coverage is therefore essential for reducing the health and economic impacts of the epidemic. Moreover, booster doses or vaccines with updated formulations may be necessary in the face of new variants. Public health authorities throughout the world have sought effective strategies to increase vaccine uptake, especially among the hesitant or procrastinating.

In response to this challenge, various local or national governments have introduced proof of vaccination mandates or certificates, [5–7], which allow vaccinated persons to attend non-essential sports or social settings and events such as concerts, stadiums, museums, restaurants, bars, etc. The goal of these policies is twofold: to provide incentives for immunization and to reduce viral transmission in risky indoor or crowded settings.

We evaluate and quantify the effect of proof of vaccination mandates on first-dose vaccine uptake in the ten Canadian provinces and three European countries (France, Italy and Germany). Some mandates accept a recent negative or past positive COVID test result as a substitute of vaccination or allow businesses to opt out if they abide by additional restrictions. Among the jurisdictions we consider, the mandates in France, Italy, Germany, Alberta and Saskatchewan allowed such options during the studied period. We chose these locations since they have similar economies, demographics and vaccine access, and all announced and implemented mandates in July–October 2021 (see Supplementary Table 1), a period with minimal binding vaccine supply or access constraints and a high base first-dose vaccination rate (above 60% of those eligible in the three countries and above 80% in Canada at the time of the mandate announcements, see Supplementary Table 2). Hence, we evaluate the mandates’ impact on people, such as the vaccine hesitant, that have remained unvaccinated for weeks or months after immunization was available to them (see Supplementary Table 3).

While, given past evidence from HPV, Tdap or hepatitis A immunizations [8–10], requiring proof of vaccination is expected to raise vaccine uptake, the magnitude and speed of the increase are hard to predict: they depend on the relative importance of the factors leading to delay or hesitancy, e.g., lack of social or economic incentives, misinformation, or entrenched political or religious beliefs. We use first doses as the main outcome in our statistical analysis because they most directly reflect the decision to be immunized.

Fig. 1 plots the weekly vaccine first doses administered in the four most populous Canadian provinces and four European countries. All except Spain introduced a province-or country-wide mandate in the studied period. We observe a sizable boost in vaccine uptake after the mandate announcement (the dashed vertical line) in all four provinces and in France, Italy and Germany, often in contrast to a sharp decline in the pre-announcement weeks. In France, daily first-dose appointments also show a striking surge on the day after the mandate announcement (see Extended Data Fig. 2). In contrast, Spain exhibits a steady decrease in weekly first doses over the displayed period.

**Fig. 1.**
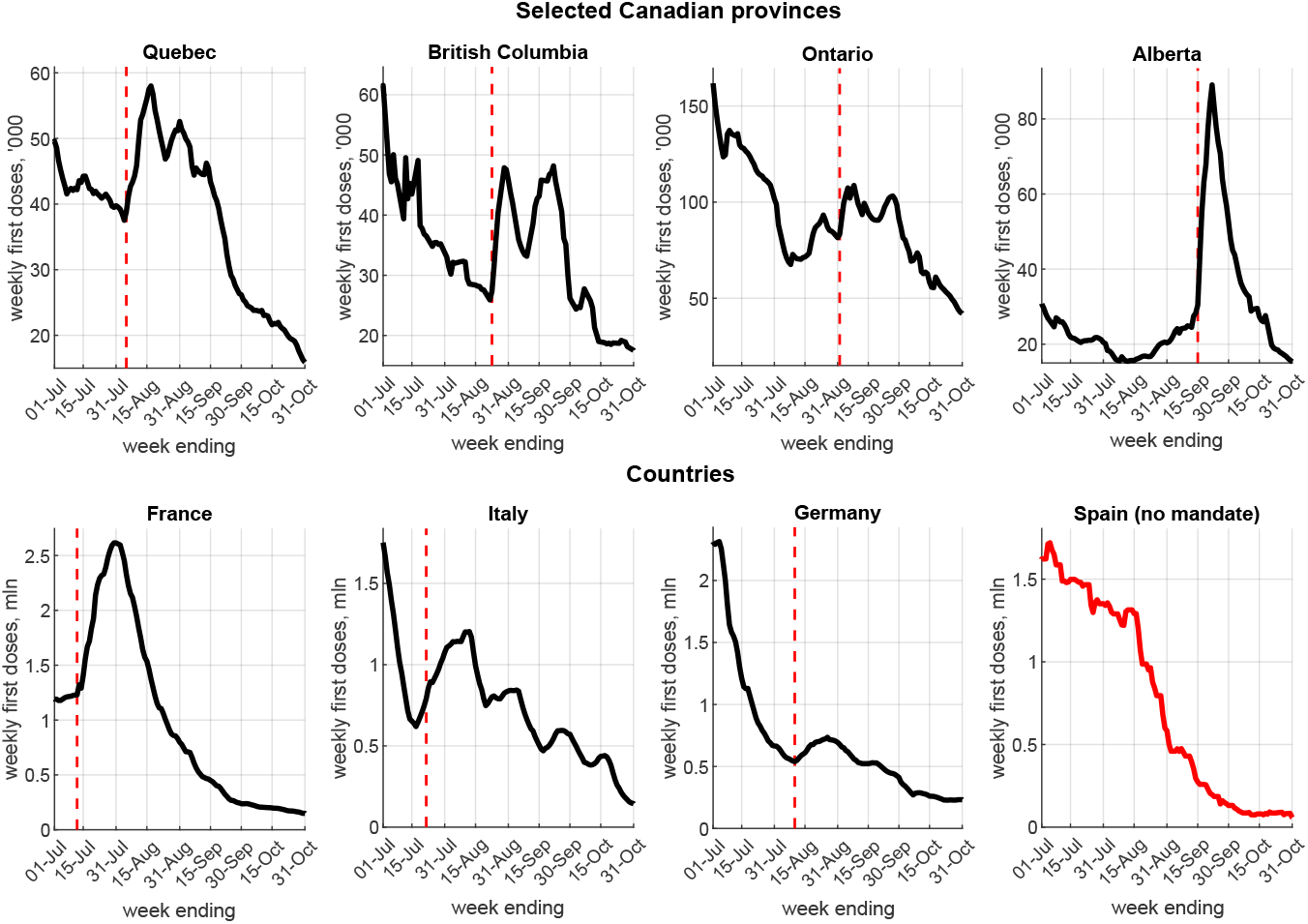
Vaccination mandates and first dose uptake. Notes: The figure plots the weekly first doses of COVID-19 vaccines administered for dates *t* −*6* to *t*, where *t* is the date on the horizontal axis. All displayed dates refer to 2021. The vertical dashed lines denote the proof of vaccination mandate announcement dates (for countries, this is the date of a *national* mandate). Spain had not announced a national proof of vaccination mandate as of October 31, 2021. We show the four most populated Canadian provinces totalling about 87% of Canada’s population (see Extended Data Fig. 3 and 4 for all ten provinces). Alberta also had a $100 debit card incentive for doses received between Sep. 3 and Oct. 14, 2021.

Motivated by this evidence, we address two important policy-relevant questions. We first estimate the magnitude of the increase in first dose vaccinations after a mandate announcement, controlling for other possible factors. Second, we evaluate how long these vaccination gains persist and the cumulative effect on vaccine uptake. We do not address ethical considerations in this paper, [11]. Our goal is to assess the mandates’ effectiveness purely in terms of raising vaccine uptake, which can then be weighed against various political or enforcement costs and compared to other policies, such as financial incentives (cash, gift cards, lotteries), [12–17], or behavioural nudges (e.g., messages from experts, appointment reminders), [12, 18–20], for which mixed or negative results have been reported.

To quantify the mandates’ impact on vaccine uptake, we use Canadian provincial data in a difference-in-differences (DID) identification strategy based on the time variation in mandate announcement dates across different geographic units in the same country, Canada (from August 5, 2021 in Quebec to September 21, 2021 in Prince Edward Island). In Canada, the provinces are separate jurisdictions with extensive powers over health policy, while the vaccines are procured by the federal government and allocated in proportion to provincial population. In contrast, the French, Italian or German mandates apply at the national level, which makes it more challenging to separate their effect from that of time trends or other concurrent events or policies.

We estimate a behavioural model in which the decision to receive a COVID-19 vaccine, measured by new weekly first doses, is affected by the policy setting (whether a proof of vaccination mandate has been announced) and the current COVID-19 epidemiological and public health conditions (‘information’), proxied by weekly cases and deaths, [21, 22]. We control for other potential confounding variables and unobserved heterogeneity with time and location (i.e., province) fixed effects (see Methods for details).

We complement and extend the DID results for Canada with a structural-break and time-series analysis, which allows us to study the mandates’ impact on vaccine uptake over a longer period. We obtain individual policy impact estimates for each Canadian province and for France, Italy and Germany, and identify potential factors contributing to the heterogeneity of the estimates, including the time between the mandate announcement and implementation and the fraction of the population that is already vaccinated at the time of announcement. We then use the time-series estimates in counterfactual simulations and compute the cumulative gains in vaccine uptake following the mandates over our study period ending October 31, 2021, relative to a hypothetical scenario in absence of mandates.

## Results

Fig. 2 plots the raw-data time profile of weekly first doses after a proof of vaccination mandate announcement, with doses in the week ending on the announcement date normalized to 100. The figure shows that weekly first doses in the Canadian provinces and in France, Italy and Germany grow quickly, reach a peak one to five weeks after the announcement date and then decrease, as in the pre-announcement trend in most locations (see Extended Data Fig. 3). New Brunswick, Newfoundland, Alberta, Nova Scotia, Saskatchewan and France registered increases in vaccine uptake of over 100% relative to the pre-announcement week. In other locations, e.g., Quebec, Ontario, Manitoba and Germany the observed increase is more moderate, less than 50%.

**Fig. 2.**
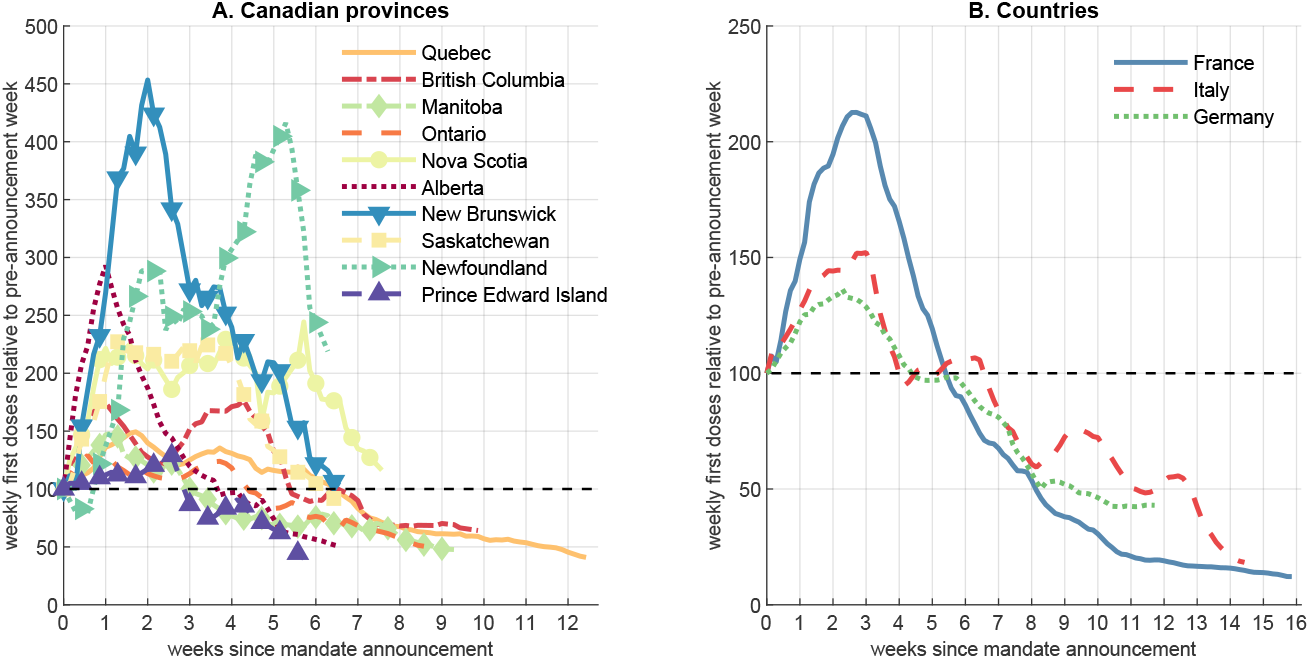
First doses after mandate announcement. Notes: The figure plots the weekly administered first doses of COVID-19 vaccine for all dates after the mandate announcement against the number of weeks since the respective announcement date (denoted by 0 on the horizontal axis), as of Oct. 31, 2021. The weekly first doses for the week just prior to the mandate announcement are normalized to 100 for each respective province (panel A) or country (panel B).

### Canadian provinces – difference-in-differences analysis

We use a difference-in-differences (DID) statistical method, the Sun and Abraham [23] treatment effect heterogeneity robust estimator (see Methods), to study the average effect of proof of vaccination mandates in Canada.

In Fig. 3, we plot results from an event study analysis of weekly first doses administered in the ten Canadian provinces, from six weeks prior to five or more weeks after the announcement of a mandate. We use as control group (latest treated) the last five provinces to announce a mandate (Alberta, New Brunswick, Saskatchewan, Newfoundland and Prince Edward Island). We chose this control group as the latest in time representative set of provinces containing a mix of smaller and larger provinces by population. This choice implies June 15, 2021 to September 14, 2021 (the day before mandate announcement in Alberta and New Brunswick, see Supplementary Table 1) as our baseline DID time period. We present results with different control groups corresponding to earlier or later sample end dates in Supplementary Table 4.

**Fig. 3.**
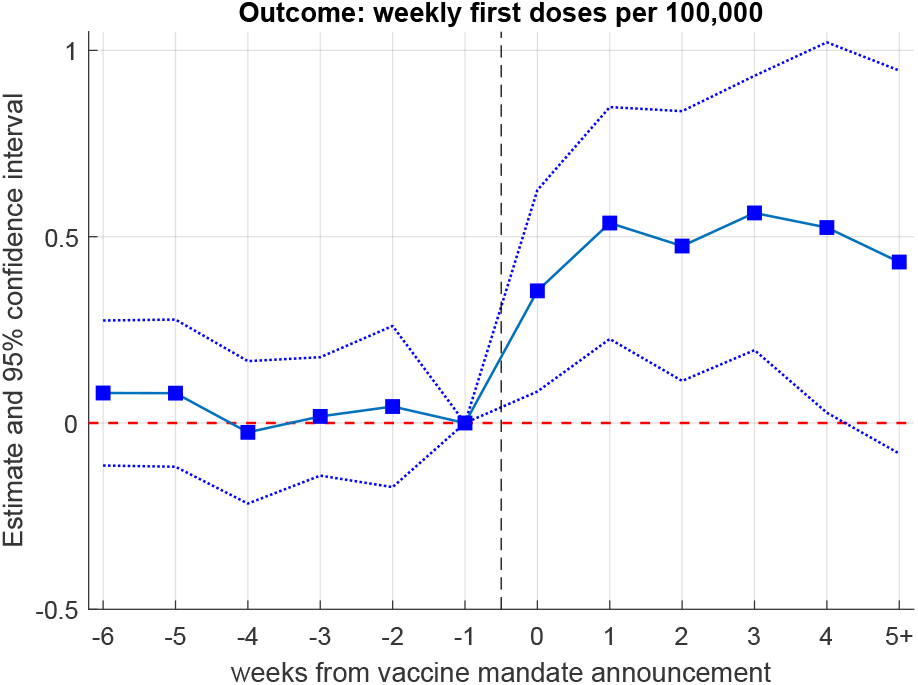
Canadian provinces – event study. Notes: Sun and Abraham [23] treatment effect heterogeneity robust estimates, see Methods. Sample period: June 15 to September 14, 2021 using Alberta, New Brunswick, Saskatchewan, Prince Edward Island and Newfoundland as the control group (latest treated). The outcome variable, *V*_*it*_ is log weekly first doses per 100,000 people administered for dates *t* − 6 to *t* inclusive. The figure plots the estimates (denoted by squares) from a variant of equation (1) where the mandate announcement variable *P*_*it*_ is replaced by the interaction of being in the ‘treatment’ group (announced mandate) with a series of dummies for each week ranging from 6 weeks before (*T* = − 6) to 5 or more weeks after the announcement (*T* = 5), where *T* = 0 denotes the week starting at the announcement date. The reference point is one week before the announcement (*T* = –1). The dotted lines correspond to 95% confidence intervals.

Fig. 3 shows a lack of mandate-associated pre-trend in the data – the DID estimates before the mandate announcement are statistically indistinguishable from zero. This addresses the potential endogeneity concern (parallel trends assumption) that provinces that announced a mandate may have had a different trend in first-dose vaccinations than provinces that did not announce a mandate. Second, the impact of the mandate announcement on first-dose vaccine uptake is realized relatively quickly and is large in magnitude — an increase of 42% (35 log points, *p* = 0.015, 95% CI 9–62) in new weekly doses in the first post-announcement week and 71% (54 log points, *p* = 0.001, 95% CI 21–86) in the second week, each relative to one week before the announcement. The observed quick increase in uptake mitigates possible concerns that vaccine supply or scheduling constraints may be affecting our results. Third, the policy effect persists over the 6-week post-announcement period we analyze (*T* = 0 to *T* = 5). Unfortunately, data limitations (the timing of the announcements) and the need for a not-yet-treated control group in the DID method do not allow us to investigate longer horizons.

Table 1 displays DID estimates of the mandate effect on first-dose uptake, relative to the pre-mandate period, controlling for information (cases and deaths) and time and location fixed effects. Columns (1) and (2) show that a mandate announcement is associated with an average increase of about 66 percent (50.6 log points, *p* = 0.001, 95% CI 25-77 in column 2) in weekly first doses. In column (3), we report estimates where the mandate policy variable is decomposed into six binary variables (one for each week after the announcement date) to account for dynamic effects. The results indicate a sharp increase in weekly first doses of 43 percent (36 log points, *p* = 0.005, 95% CI 14–58) in the week beginning at the announcement date (week 0). The increase is sustained throughout the post-announcement sample period, as all estimates in column (3) are positive (*p <* 0.06) and larger than the week 0 estimate. The DID analysis thus yields no evidence that short-term intertemporal substitution (which would be manifested as negative estimates in the later weeks) is the mechanism behind the observed boost in first-dose vaccinations in the studied period.

**Table 1.**
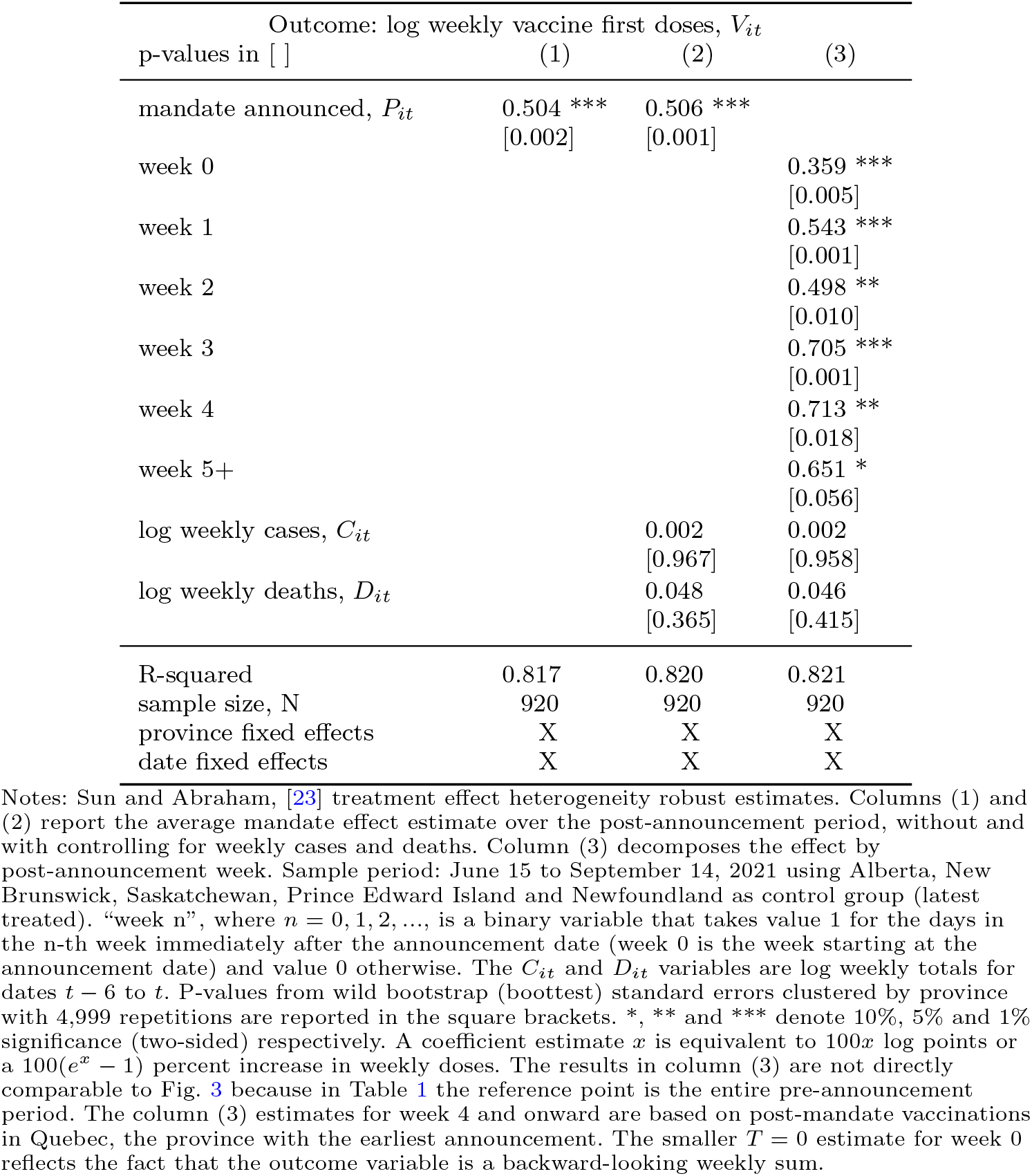
Canadian provinces – difference-in-differences estimates

| Outcome: log weekly vaccine first doses, $V_{it}$ | | | |
| --- | --- | --- | --- |
| p-values in [ ] | (1) | (2) | (3) |
| mandate announced, $P_{it}$ | 0.504 ***<br>[0.002] | 0.506 ***<br>[0.001] | |
| week 0 |  |  | 0.359 ***<br>[0.005] |
| week 1 |  |  | 0.543 ***<br>[0.001] |
| week 2 |  |  | 0.498 **<br>[0.010] |
| week 3 |  |  | 0.705 ***<br>[0.001] |
| week 4 |  |  | 0.713 **<br>[0.018] |
| week 5+ |  |  | 0.651 *<br>[0.056] |
| log weekly cases, $C_{it}$ | | 0.002<br>[0.967] | 0.002<br>[0.958] |
| log weekly deaths, $D_{it}$ | | 0.048<br>[0.365] | 0.046<br>[0.415] |
| R-squared | 0.817 | 0.820 | 0.821 |
| sample size, N | 920 | 920 | 920 |
| province fixed effects | X | X | X |
| date fixed effects | X | X | X |

We perform a range of robustness checks and sensitivity analysis on our main DID results. Our result of a more than 60% increase in weekly first doses on average over the post-announcement weeks remains robust when using alternative dates and treatment groups varying from the first three to the first nine provinces to announce a mandate (see Supplementary Table 4 and Extended Data Fig. 5). We also allow for alternative initial sample dates or lags in the policy or information variables (see Extended Data Fig. 5). Additional robustness checks include using different control variables (hospitalizations), population weights, OLS two-way fixed effects, daily data, levels data, alternative ways to compute the standard errors, and using randomized (placebo) announcement dates, see Extended Data Fig. 6 and Supplementary Tables 5 and 6 for details. A possible concern is that case or death counts may be correlated with the mandate announcements (while also affecting vaccination rates as information, for which we control). We ran event-study analyses analogous to that in Fig. 3 but using weekly cases or deaths as the outcome and find that the estimates before the announcements are statistically indistinguishable from zero. This suggests an absence of significant differential pre-trends in cases or deaths across the treated and control provinces.

Most provincial proof of vaccination mandates required two doses to be considered adequately vaccinated with a vaccine offered in Canada during the study period: Pfizer (Comirnaty), Moderna (SpikeVax) or AstraZeneca (Vaxzevria); there were limited exceptions in Quebec (a prior infection could count as first dose) and British Columbia (only one dose was required between Sep. 13 and Oct. 23, 2021). In Supplementary Table 7, we use log of weekly second doses as the outcome. We do not find a statistically significant effect of mandate announcements on second dose uptake in our sample period, consistent with second doses being more evenly spread out over time. Extended Data Fig. 7 also shows that, unlike the sharp boost in first doses, there are only small or gradual post-mandate increases in second doses in some of the provinces. One possible explanation for the lack of large increases in second doses a few weeks after the spikes in first doses is that mandates may have encouraged some people that already had their first doses to obtain their second dose sooner, thus shifting some second doses forward and dampening the lagged effect.

We also do not find statistically significant effect on first-dose uptake associated with the mandates’ implementation dates additional to the announcement date effect; see Supplementary Information for details.

### Time-series analysis

We complement and extend the DID panel-data results with a structural break and time-series analysis using the ‘interrupted time-series analysis’ (ITSA) method [24], which models the relationship between the outcome (weekly first doses) and the policy variable (mandate announcement) and controls for time trends, lagged outcomes and information (weekly cases and deaths), see Methods for details. This allows us to estimate the mandates’ longer-term impact on vaccine uptake separately for each of the ten Canadian provinces, as well as for France, Italy and Germany, using all the data up to October 31, 2021.

We first test for the presence of a structural break at the announcement date and find that we can reject the null hypothesis of no break for each of the provinces and countries; see Supplementary Table 8 and Methods for details. In Table 2, we report the time-series estimates for each country and province in our data. In columns (1)-(3), we report the estimated coefficients on the mandate announcement variable, 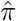 and on two time trends: a linear daily time trend (estimate 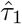) and an ‘interaction’ time trend reflecting the post-mandate trend slope change (estimate 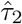). Since the outcome and information variables are level variables, in column (4), we use the augmented Dickey-Fuller (ADF) unit root test to test whether the residuals are stationary, to avoid a spurious regression. We reject the null hypothesis that the residual contains a unit root at the 95% confidence level for all locations except Newfoundland, where we can reject the null at the 90% level.

**Table 2.** Time series estimates

| Outcome: log weekly vaccine first doses |  |  |  |  |
| --- | --- | --- | --- | --- |
| p-values in [ ] | policy $\hat{\pi}$<br>(1) | time<br>trend $\hat{\tau}_1$<br>(2) | trend<br>change $\hat{\tau}_2$<br>(3) | ADF test<br>(4) |
| **Countries** |  |  |  |  |
| France | 0.154 ***<br>[0.004] | -0.018 ***<br>[0.000] | 0.003<br>[0.229] | -3.970<br>[0.002] |
| Italy | 1.025 ***<br>[0.000] | -0.029 ***<br>[0.001] | 0.005<br>[0.556] | -3.365<br>[0.012] |
| Germany | 0.438 ***<br>[0.000] | -0.017 ***<br>[0.000] | 0.006 **<br>[0.012] | -3.235<br>[0.018] |
| **Canadian provinces** |  |  |  |  |
| Quebec | 0.064<br>[0.647] | -0.029 **<br>[0.010] | 0.017 *<br>[0.079] | -5.192<br>[0.000] |
| British Columbia | 0.734 ***<br>[0.000] | -0.013 ***<br>[0.008] | -0.004<br>[0.400] | -4.587<br>[0.000] |
| Manitoba | 0.757 ***<br>[0.000] | -0.034 ***<br>[0.000] | 0.012 **<br>[0.014] | -4.735<br>[0.000] |
| Ontario | 0.293 ***<br>[0.007] | -0.011 **<br>[0.033] | 0.002<br>[0.739] | -4.166<br>[0.001] |
| Nova Scotia | 0.595 ***<br>[0.002] | -0.025 ***<br>[0.000] | 0.021 **<br>[0.021] | -4.698<br>[0.000] |
| Alberta | 1.455 ***<br>[0.000] | -0.015 ***<br>[0.000] | -0.024 ***<br>[0.000] | -3.372<br>[0.012] |
| New Brunswick | 1.313 ***<br>[0.000] | -0.011 **<br>[0.019] | -0.017 ***<br>[0.004] | -4.416<br>[0.000] |
| Saskatchewan | 0.898 ***<br>[0.000] | -0.008 ***<br>[0.001] | -0.013 **<br>[0.013] | -3.800<br>[0.003] |
| Newfoundland | 0.410 *<br>[0.075] | -0.007<br>[0.241] | 0.005<br>[0.742] | -2.689<br>[0.076] |
| Prince Edward Island | 0.378 **<br>[0.010] | -0.002<br>[0.565] | -0.014 **<br>[0.019] | -2.904<br>[0.045] |
| sample size for each row | 139 | 139 | 139 | 139 |

Notes: Time period – June 15 to October 31, 2021. Each row is a separate time-series regression as specified in (2). All rows include 7-day and 14-day lags of the outcome variable and log weekly deaths $D_t$ and log weekly cases $C_{it}$ as information, $I_t$ . Column (1) reports the estimate $\hat{\pi}$ on the mandate announcement policy variable $P_t$ in equation (2). Column (2) reports the estimate $\hat{\tau}_1$ on the linear time trend $T_t$ . Column (3) reports the estimate $\hat{\tau}_2$ of the post-announcement trend slope change. P-values computed using Newey-West heteroskedasticity and autocorrelation robust (HAC) standard errors with 3 lags are in the square brackets. \*, \*\* and \*\*\* denote 10%, 5% and 1% significance (two-sided) respectively. Column (4) reports augmented Dickey-Fuller (ADF) one-sided test statistics and p-values using 13 lags chosen as the integer part of $12(T/100)^{1/4}$ ; see [25].

The estimates, 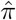 for the initial rise in weekly first doses after mandate announcement in Table 2 are large and statistically significantly positive for all three countries: 17% (15.4 log points, *p* = 0.004, 95% CI 5.0–25.8) for France, 179% (103 log points, *p <* 0.001, 95% CI 53–152) for Italy and 55% for Germany (43.8 log points, *p <* 0.001, 95% CI 31.4–56.1), relative to the respective pre-mandate trends. A comparison of Fig. 1 and Extended Data Fig. 2 suggests that the relatively low French estimate may reflect a lag between appointment booking and vaccine administration. We also obtain statistically significantly positive and large estimates of the increase in weekly first doses after mandate announcement, 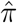, for all provinces except Quebec. The estimate for Quebec is 0.389 (*p <* 0.001, 95% CI 0.265–0.514) and statistically significantly different from zero when we use deaths and hospitalizations as information; see Supplementary Table 9. The estimated mandate effect varies across the provinces: e.g., a 34% (29 log points, *p* = 0.007, 95% CI 8.2–50.5) initial increase in Ontario vs. 326% (145 log points, *p <* 0.001, 95% CI 110– 181) in Alberta. The range of the provincial estimates is consistent with our DID estimate in Table 1 for the average mandate effect for Canada; however, the two estimation methods are not directly comparable.

The estimated mandate effect on vaccine uptake tends to be larger for provinces experiencing a surge in cases at the same time (for which we control), namely Alberta, Saskatchewan and New Brunswick; see Extended Data Fig. 3. These provinces also announced their mandates relatively late (mid-September) and set a relatively short time interval between mandate announcement and implementation (see Supplementary Table 1). A larger fraction of eligible unvaccinated people at the time of mandate announcement is also positively associated with a larger policy effect estimate. We illustrate these associations in Extended Data Fig. 8.

The baseline time trend in first doses is downward sloping 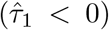 in all countries and provinces (although not statistically significant in the two smallest provinces, Newfoundland and Prince Edward Island) and indicates a steady decrease in new vaccinations of 0.8% to 3.4% per day in the studied period in absence of mandate. The post-mandate announcement trend in first doses turns less steep in the three countries (the estimate 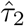 is positive, although only statistically significant for Germany), which suggests a lack of net intertemporal substitution as of October 31. On the other hand, the estimates 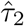 are statistically significantly negative for Alberta, New Brunswick, Saskatchewan and Prince Edward Island, which is consistent with the mandate effect on new first doses diminishing over time in these provinces.

In Supplementary Table 9, we report additional results from two alternative time-series specifications: using weekly deaths and average hospitalizations as information control variables or using a binary, instead of weekly-averaged, policy variable. The results are very similar to those in Table 2 and confirm the large and positive estimated effect of mandate announcements on first dose vaccine uptake. We provide further details on the time-series model fit and out-of-sample projections in Supplementary Information.

### Counterfactuals

We use our estimates of the mandate effect on vaccine uptake from Table 2 to compute the cumulative increase in new vaccinations for each province and for France, Italy and Germany, relative to the counterfactual scenario of absence of mandate. Counterfactual weekly and cumulative doses are computed as explained in Methods. We report the vaccinations gains both in levels (mln doses) and in percentage points relative to the vaccination rate at the mandate announcement date.

We find large cumulative increases in the first-dose vaccination rate for all provinces relative to in the absence of mandate: from 1.9 p.p. (90% CI -0.3– 3.0) in Ontario to 5 p.p. (90% CI 4.0–5.6) in Saskatchewan and 5 p.p. (90% CI 3.9–5.8) in New Brunswick, with all other provinces in between, as of October 31, 2021; see Fig. 4 and Fig. 5. These estimated total gains in uptake add up to 2.9 p.p. (90% CI 1.3–3.8) of the eligible population or 979,000 new first doses (90% CI 425,000–1,266,000) for Canada as a whole; see Supplementary Table 10. This is a significant increase in vaccine uptake considering the relatively short period in which it was achieved (within 6 to 10 weeks of the mandate announcement for most provinces; 13 weeks for Quebec) and the very high pre-mandate first-dose vaccination rate in Canada (over 80% on average at the time of the mandate announcements, see Supplementary Table 2). For example, Saskatchewan’s 5 p.p. estimated gain amounts to about one quarter of all remaining unvaccinated eligible people in the province as of the mandate announcement date (Sep. 16, 2021).

**Fig. 4.**
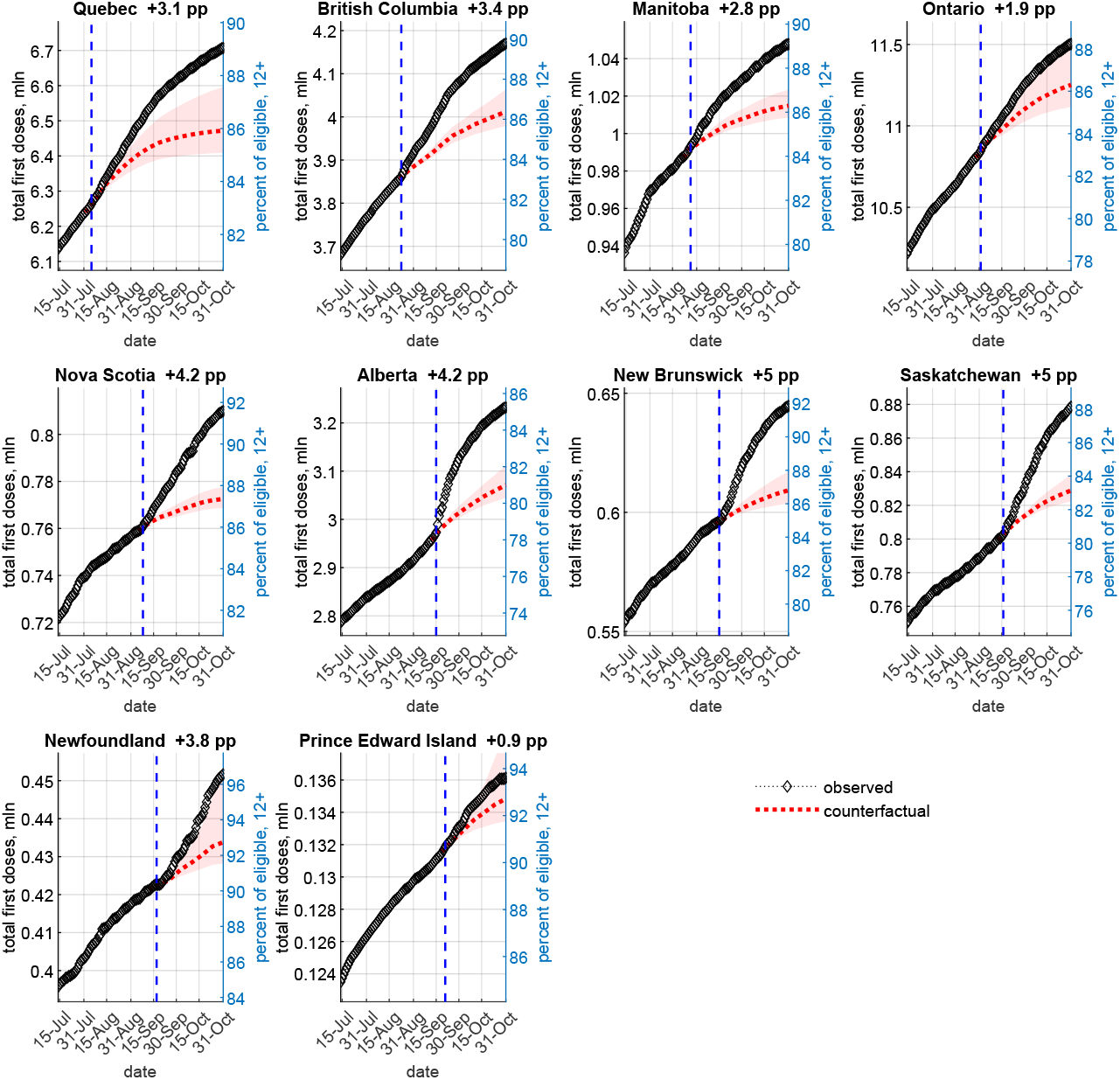
Canadian provinces – observed vs. no-mandate counterfactual first doses as of October 31, 2021 (time-series estimates) Notes: The figure plots the observed (diamonds) and estimated mean counterfactual (dotted line) cumulative first doses for each province by date. All displayed dates refer to 2021. The counterfactuals are computed using the estimates in Table 2. The shaded areas denote 5–95 percentile confidence bands computed computed using 1000 draws from the estimated asymptotic joint distribution of the parameters in (3). The vertical dashed line denotes the mandate announcement date. The number next to each province name indicates the mean estimated percentage point increase in first doses relative to the no-mandate counterfactual, as of October 31, 2021.

**Fig. 5.**
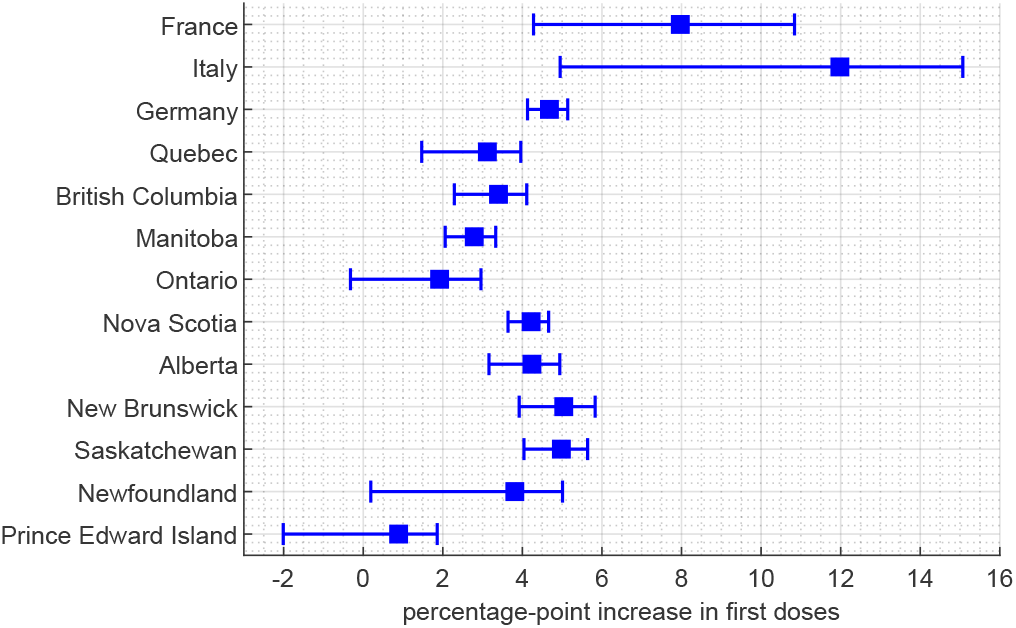
Counterfactuals – percentage-point increase in vaccine uptake. Note: The figure plots the total estimated percentage-point increase (mean and 90% confidence interval) in the first dose vaccination rate relative to the no-mandate counterfactual, as of October 31, 2021.

We do not find evidence of net intertemporal substitution (decrease in the cumulative first-dose gains from pulling vaccinations forward in time) as of October 31, 2021, except in PEI. However, the strongly negative 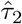 estimates in Table 2 and the trends in Extended Data Fig. 9 indicate that Alberta, New Brunswick and Saskatchewan are projected to exhibit net intertemporal substitution after the end of October 2021. That said, given the flattening of the counterfactual curves on Fig. 4, the cumulative gains are likely to remain significant.

In Extended Data Fig. 10, we also plot a counterfactual computed using the DID estimate for Canada from Table 1 (i.e., using the same policy estimate for all provinces). We find that, as of the DID sample end date of September 14, 2021, the five Canadian provinces that announced mandates by that date have benefited from 287,000 additional first doses (90% CI 239,000–333,000) or a vaccination rate increase of 0.9 p.p. (90% CI 0.7–1.0) for the eligible population, relative to the no-mandate counterfactual.

We also estimate large increases in first-dose vaccinations relative to the no-mandate counterfactual in the three countries: 8 p.p. (90% CI 4.3–10.8) or 4.59 mln (90% CI 2.47–6.25) additional first doses in France, 12 p.p. (90% CI 5.0–15.1) or 6.48 mln (90% CI 2.67–8.14) doses in Italy, and 4.7 p.p. (90% CI 4.1–5.1) or 3.47 mln (90% CI 3.06–3.81) doses in Germany as of October 31, 2021 using our main Table 2 specification, see Fig. 5, Fig. 6 and Supplementary Table 10. These estimated gains are larger than those for Canada, possibly because of the earlier mandates in these countries or the lower starting vaccination rates, and they may partly reflect expanding the scope of the initial mandates, e.g., to inter-regional travel and employment in September and October in Italy.

**Fig. 6.**
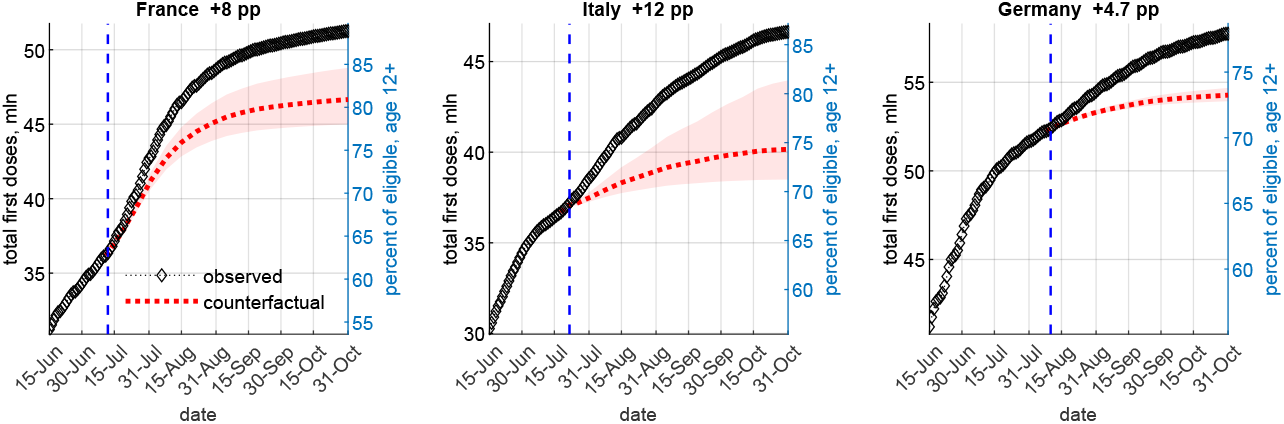
Countries – observed vs. no-mandate counterfactual first doses. Notes: The figure plots the observed (diamonds) and estimated mean counterfactual (dotted line) cumulative first doses for each country by date. All displayed dates refer to 2021. The counterfactuals are computed using the estimates from Table 2. The shaded areas denote 5–95 percentile confidence bands computed using 1000 draws from the estimated asymptotic joint distribution of the parameters in (3). The vertical dashed line denotes the mandate announcement date. The number next to each country name indicates the mean estimated percentage point increase in first doses relative to the no-mandate counterfactual, as of October 31, 2021.

The counterfactuals assume that all explanatory variables except the mandate announcement (e.g., cases, deaths or time trends) remain fixed at their observed values and that the model parameters remain stable. These assumptions are more plausible over relatively short periods. Hence, these simulations should be interpreted primarily as an illustration of the estimated impact of the mandates on vaccine uptake rather than policy guidance.

## Discussion

We find that government-mandated proof of vaccination requirements or certificates have sizable and statistically significant impact on COVID-19 vaccine uptake, with large observed increase in first-dose vaccinations in the first several weeks after mandate announcement and lasting cumulative gains relative to the pre-announcement trend. This includes robust difference-in-differences evidence using the variation in the timing of mandate announcements within the same country, Canada. Our results are of similar magnitude and consistent with the findings of other authors, using different data samples and methodologies: [26] use a synthetic control approach and estimate large increases in vaccinations in France (8.6 mln), Italy (4 mln) and Israel (2.1 mln) from 20 days prior to 40 days after COVID-19 certification mandate implementation, while [27] compute counterfactuals based on an innovation diffusion model of vaccine uptake and attribute vaccination rate increases of 13 p.p. for France, 6.2 p.p. for Germany and 9.7 p.p. for Italy to the announcement of COVID-19 certificate requirements, with associated additional gains from averted deaths and GDP losses.

The estimated mandate effect on uptake varies across the Canadian provinces, with the timing of announcement, implementation and the percentage unvaccinated playing a role. The estimated impact also differs across France, Italy and Germany. Further research on understanding this heterogeneity and on other potentially important factors (e.g., the role of government communication or the media, the degree of political polarization, the amount of social trust) can complement our study.

The unambiguous and large increases in vaccine uptake we find compares favourably to the mixed evidence from using financial incentives (cash, gift cards, lotteries), [12–17], or behavioural nudges, [12, 18–20]. Financial incentives for vaccination have been criticized for the optics of putting a low dollar value on being vaccinated compared to the social benefits, because of the perceived unfairness in rewarding people who delayed their vaccination, or because of potential moral hazard problems (e.g., expecting future payments). Financial incentives may even have the perverse effect of validating vaccine concerns among unvaccinated individuals; see [12]. Others [28] have argued that, given the already high vaccination rates in developed countries, behavioural nudges may not be very effective, which is consistent with the findings of [12] and [13]. On the other hand, vaccination mandates have been controversial, as some people perceive them as restrictions on personal freedom. This can affect compliance and increase both the direct implementation and enforcement costs, as well as the political costs of introducing a mandate.

In terms of external validity, in 2020, [29] conducted a survey on vaccine hesitancy across 19 countries comprising around 55% of the world population. The participants were asked: “If a COVID-19 vaccine is proven safe and effective and is available, will you take it?” Canada, Italy and Germany placed around the middle in the self-reported vaccine hesitancy rate (29% to 31%), while France had a higher hesitancy rate (41%). In this regard, our results on the large impact on new first dose vaccinations in these countries can be useful to public health authorities in other places looking for an effective strategy to increase vaccine uptake.

We conclude by listing some limitations and areas for further research. We have abstracted from assessing the mandates’ impact on health outcomes (cases, hospitalizations or deaths). However, vaccine effectiveness estimates from the medical literature can be used to study this further, ideally controlling for possible changes in behaviour. Our focus is on vaccination and certification requirements for non-essential settings although some locations, e.g., Italy, expanded the mandates to inter-regional travel or employment and other countries (Greece, Austria) have proposed even broader mandates, which we do not analyze. For data limitation reasons, we also could not study the effect by age group as in [26] or the possible role of past personal sickness outcomes as in [30]. See also Methods for limitations and required assumptions related to the time-series analysis and counterfactuals and our ways of tackling them.

Naturally, a full cost-benefit analysis of proof of vaccination mandates is beyond the scope of this paper. In particular, the costs of imposing and enforcing the mandates – economic, political, or personal – are very hard to estimate, as is the social value of vaccinating an additional person, [31, 32]. One component of the latter is avoided healthcare costs, e.g., [14] estimate that the Ohio vaccine lottery saved the state USD 66 mln in ICU costs. Our results are a step toward quantifying the benefits of proof of vaccination requirements.

## Methods

### Data and definitions

We use data on COVID-19 vaccination numbers, cases and deaths for all ten Canadian provinces and for France, Italy, Germany and Spain; see Supplementary Table 11 for details. We collected the Canadian data from the official provincial dashboards or equivalent sources. We use the Our World In Data (OWID) dataset for country data. Announcement and implementation dates of the proof of vaccination mandates or certificates were collected from government websites and major newspapers; see Supplementary Table 1.

The main variables used in our statistical analysis are defined next. Everywhere, *i* denotes province or country and *t* denotes time measured in days (date). We aggregate the data on vaccinations, cases and deaths on a weekly basis (totals for the week ending on date *t*, i.e., dates *t* − 6 to *t*) to reduce the influence of day-of-the-week effects or reporting artifacts (e.g., lumping week-end data in Monday’s report; in the latter case, we distribute the reported total equally over the affected dates).

- Outcome, *V*_*it*_. The main outcome variable is the logarithm of administered vaccine first doses per 100,000 people, for the week ending at *t* (dates *t* − 6 to *t*). We use first doses, as they most directly reflect the impact of the mandates on the intent to be immunized and avoid potential issues related to second dose scheduling or availability. In Supplementary Table 7, we also report results using second doses as the outcome. All COVID-19 vaccines used in Canada during the study period, namely Pfizer (Comirnaty), Moderna (SpikeVax) and AstraZeneca (Vaxzevria), were originally considered two-dose. Using the logarithm of weekly first doses allows us to interpret the regression coefficients as percentage changes; moreover, the estimates are invariant to normalization, e.g., by population (subsumed in the regression constant or fixed effects). We thus use “log weekly first doses” for simplicity throughout the text, except where the actual scale is important.
- Policy, *P*_*it*_. Let 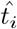 be the mandate announcement date in jurisdiction *i*. We construct a binary policy variable *P*_*it*_ equal to 1 for all post-announcement dates *t* ≥ 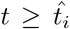 and equal to 0 for all 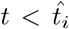. Proof of vaccination mandates were announced in all ten Canadian provinces over the period Aug. 5, 2021 to Sep. 21, 2021; see Extended Data Fig. 4 and Supplementary Tables 1 and 12. Last to announce were the four Atlantic provinces, which had the lowest per capita case rates in August 2021, and Alberta and Saskatchewan, which had the highest per capita case rates in August 2021; see Extended Data Fig. 3. France, Italy and Germany also introduced proof of vaccination certificates in July – August 2021; see Supplementary Table 1.
- Information, *I*_*it*_. We include control variables related to the concurrent COVID-19 epidemiological situation, specifically log of weekly cases, *C*_*it*_, and log of weekly deaths, *D*_*it*_, for the week ending at date *t* (dates *t* − 6 to *t*). We refer to these variables jointly as ‘information’, *I*_*it*_ (see [33, 34]) since they can inform a person’s COVID-19 exposure risk assessment and/or decision to be vaccinated, e.g., as shown in [21, 22]. Another possible information variable is hospitalizations. However, it is strongly correlated with COVID-19 cases and deaths so all three cannot be included at the same time (see Supplementary Tables 5 and 9 for results using deaths and hospitalizations as information). To address zero weekly values, which sometimes occur in the smaller provinces for deaths or cases (4.4% of all observations for cases and 10.7% for deaths), we replace log(0) with −1, as in [33].
- Other controls, *W*_*it*_. We include province fixed effects and date fixed effects in our difference-in-differences analysis. The province fixed effects account for any time-invariant province characteristics such as sentiment towards vaccination, age structure, education, political alignment, etc. The date fixed effects control for national trends or events, e.g., public messaging, vaccine-related international travel regulations, or campaigning for the 2021 federal election. In the time-series analysis, we control for time trends.
- Time period. We use the period June 15, 2021 to October 31, 2021. The start date was chosen to ensure that possible constraints in obtaining a first dose related to eligibility or supply are minimal or non-existent. This helps avoid potential bias from constrained vaccine supply affecting the pre-or post-mandate pace of vaccination. In Canada, the provinces opened registration for first-dose vaccination for any person of age 12 or older between May 10, 2021 in Alberta and May 27, 2021 in Nova Scotia; see Supplementary Table 3. First-dose availability in France, Italy and Germany was similar by mid-June, at least for the 18-plus age group. We explore different sample start dates in robustness checks; see Extended Data Fig. 5. The sample end date is based on data availability at the time of statistical analysis and writing. First doses for the 5–11 age group were not approved in the studied period.

### Difference-in-differences estimation

We estimate a behavioural model in which the decision to receive a COVID-19 vaccine, measured by new weekly first doses, is affected by the policy setting, *P*_*it*_ (whether a mandate has been announced) and current COVID-19 epidemiological information about public health conditions, *I*_*it*_, proxied by weekly cases and deaths. Based on the raw data patterns in Fig. 1 and Extended Data Fig. 3 and the absence of vaccine supply constraints in the studied period, we assume no lag between a mandate announcement and a person’s ability to receive a first vaccine dose and no information lag. We perform sensitivity analysis using alternative lags (see Extended Data Fig. 5); the results affirm our baseline choice of no lag.

In the difference-in-differences (DID) analysis, we estimate:

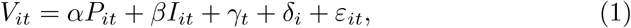

where *V*_*it*_ is log weekly first doses for location *i* and date *t, I_it_* is information, *γ*_*t*_ are date fixed effects, *δ*_*i*_ are province fixed effects, and *ε*_*it*_ is an error term. The coefficient *α* on the policy variable *P*_*it*_ captures the average effect of the mandate announcement on weekly first doses over all post-announcement dates. To capture dynamic effects of the mandates, we also estimate a version of equation (1) with *P*_*it*_ split into separate indicator variables, one for each week after the mandate announcement date; see Table 1.

To correct for the small number of clusters in the estimation since there are only ten provinces, we report “wild bootstrap” p-values (we use the Stata package *boottest* clustered by province with 4,999 repetitions, see [35, 36]). The use of clustered standard errors allows the error terms to be to be serially correlated within each province. Alternative methods for computing the standard errors are explored in Supplementary Table 6 – clustering at the province level (Stata command “cluster”), wild bootstrap standard errors clustered at the province level and wild bootstrap standard errors two-way clustered by province and date allowing for spatio-temporal correlation (Stata command “boottest”).

The recent methodological literature, [23, 37–40], has argued that the standard OLS two-way fixed effects (TWFE) estimator can be invalid in panel-data settings with staggered adoption like ours if there is heterogeneity in the treatment effect across cohorts (provinces in our data) and/or over time. The reason is that the TWFE estimate is a weighted average of many 2-by-2 DID treatment effects, where some of the weights can be negative or incorrect because of contamination from other periods.

In particular, Sun and Abraham [23] develop a difference-in-differences estimator valid under these conditions, which we use to estimate (1). The Sun and Abraham estimator uses never-treated or last-treated units as the control group and is constructed as weighted average of treatment effects for each cohort (by date of mandate announcement) and each relative time after or before the announcement (we use the Stata function *eventstudyinteract* provided by the authors). Specifically, to calculate the average treatment effect *α*, we replace *αP_it_* in (1) with

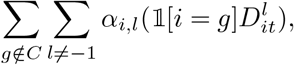

where *C* is the set of never-treated or last-treated provinces (control group), 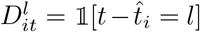 is a “relative time” indicator, and 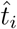 is the date of treatment for province *i*. Under parallel trends and no anticipation, [23] show that *α*_*i,l*_ is consistent for the province-time specific treatment effect. Then, the average treatment effect for each relative-time period, *α*_*l*_, is the appropriately weighted (by the sample share of each treated province in relative time *l*) average of *α*_*i,l*_ across the units *i*, and *α* in (1) is computed as the simple average of *α*_*l*_ across *l*.

The estimator requires excluding all time periods in which units in the control group are treated. Since the last province to announce a mandate was Prince Edward Island, on September 21, 2021, we can use only data until September 20, at the latest, see Supplementary Table 1. We present results for different control groups and corresponding sample periods in Supplementary Table 4. We also compare our main results with the OLS TWFE estimates in Supplementary Table 5.

### Time-series estimation

We estimate the following ‘interrupted time-series analysis’, [24], specification for each country or province (we omit the subscripts *i* for simplicity since all variables refer to the same location):

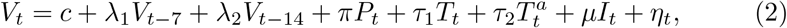

where *V*_*t*_ are log weekly first doses for the week ending at date *t, c* is a constant, *I*_*t*_ is proxy for information, analogous to its counterpart in equation (1), and *η*_*t*_ is the error term. In our baseline specification in Table 2, we construct the policy variable *P*_*t*_ as the weekly average from date *t* − 6 to *t* of the ‘mandate announced’ indicator *P*_*it*_ defined above. This is consistent with the weekly vaccination and information variables *V*_*t*_ and *I*_*t*_ and improves the fit in the time-series regressions. We also present results without weekly averaging in Supplementary Tables 9 and 10. We include 7-day and 14-day lagged values of *V*_*t*_ (instead of first and second lags) since the outcome variable *V*_*t*_ is a weekly total.

We include two time trends in equation (2): *T*_*t*_ is a linear daily time trend initialized at the sample start date *t* = 0 and 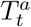 is an ‘interaction’ time trend which takes value 0 at all dates until the announcement date (inclusive) and increases by 1 for each day afterwards. The coefficients *τ*_1_ (slope) and *τ*_2_ (change in slope at the announcement date) characterize respectively the baseline (pre-announcement) time trend (with slope *τ*_1_) and the post-announcement time trend (with slope *τ*_1_ + *τ*_2_) in weekly first doses.

Standard errors and p-values are calculated using the Newey-West [41] heteroskedasticity and autocorrelation (HAC) robust estimator, with 3 lags. The lag was chosen as the closest integer to *T* ^1*/*4^, where *T* is the sample size.

The time-series approach requires stronger identification assumptions than the DID approach since there is no control group. Specifically, we need to assume exogeneity of the announcement date and that the time-series process for weekly first doses changes after the announcement only because of the policy, i.e., it would have followed the same pre-trend if no mandate had been announced. We also cannot control for fixed effects in a flexible way beyond including a constant and time trends. Under these assumptions, the coefficient on the policy variable *P*_*t*_ captures the average shift in first-dose uptake attributed to the mandate announcement. Similarly, the coefficient on the interaction time trend 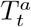 measures the slope change in the trend of first doses after the mandate announcement relative to the pre-announcement trend.

### Structural break at the announcement date

We perform a structural break Chow test, [42], for a known break point using the log weekly first doses, *V*_*t*_ in Supplementary Table 8. The presence of a break point at the mandate announcement date indicates an abrupt change or shift in the first-dose time series process. We use a bandwidth of 50 days before and 35 days after the announcement date. The unequal before-after bandwidth is chosen to reduce the size distortion of the test, since the outcome variable is a weekly sum and the error terms are serially correlated; see [43]. Columns (1) and (2) in Supplementary Table 8 use the log of weekly first doses, *V*_*t*_ while column (3) uses first-differenced weekly first doses. The differenced series is stationary, and the error terms are not serially correlated, which alleviates concerns about size distortion in the test. The power of the Chow test is weaker in this specification since the first-differenced *V*_*t*_ series is a growth rate being used to test for a level shift. The differenced series is also noisier as it captures daily fluctuations. Overall, the structural break test results show that a mandate announcement is strongly associated with a trend break in first-dose vaccine uptake in all or most locations.

### Counterfactuals

Calling 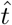 the mandate announcement date, we compute the counterfactual log weekly doses per 100,000 people, 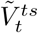 iteratively using

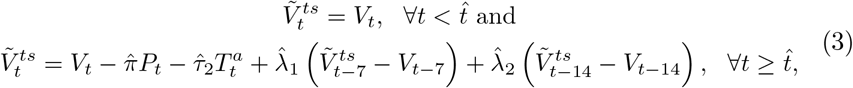

where 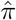 is the coefficient estimate of the mandate announcement variable *P*_*t*_ from column (1) of Table 2, 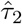 is the estimate of the interaction time trend 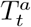 from column (2) of Table 2, and where the terms multiplied by the estimates 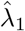 and 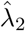 account for the the lagged values *V*_*t*_−7 and *V*_*t*_−_14_ in equation (2). The policy effect estimate 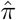, as well as 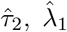 and 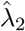 are specific for each respective country or province. To plot Extended Data Fig. 10, we compute the counterfactual mean as 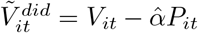 where 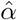 is the DID estimate on the policy variable *P*_*it*_ from Table 1, column (2).

## Data availability

All data used in this paper are publicly available at https://github.com/C19-SFU-Econ/dataV

## Data Availability

All data used in this manuscript is based on information available in public domain. Data sources and links are provided within the manuscript.

## Author contributions

All authors contributed equally to this research.

## Extended Data

**Extended Data Fig. 1.**
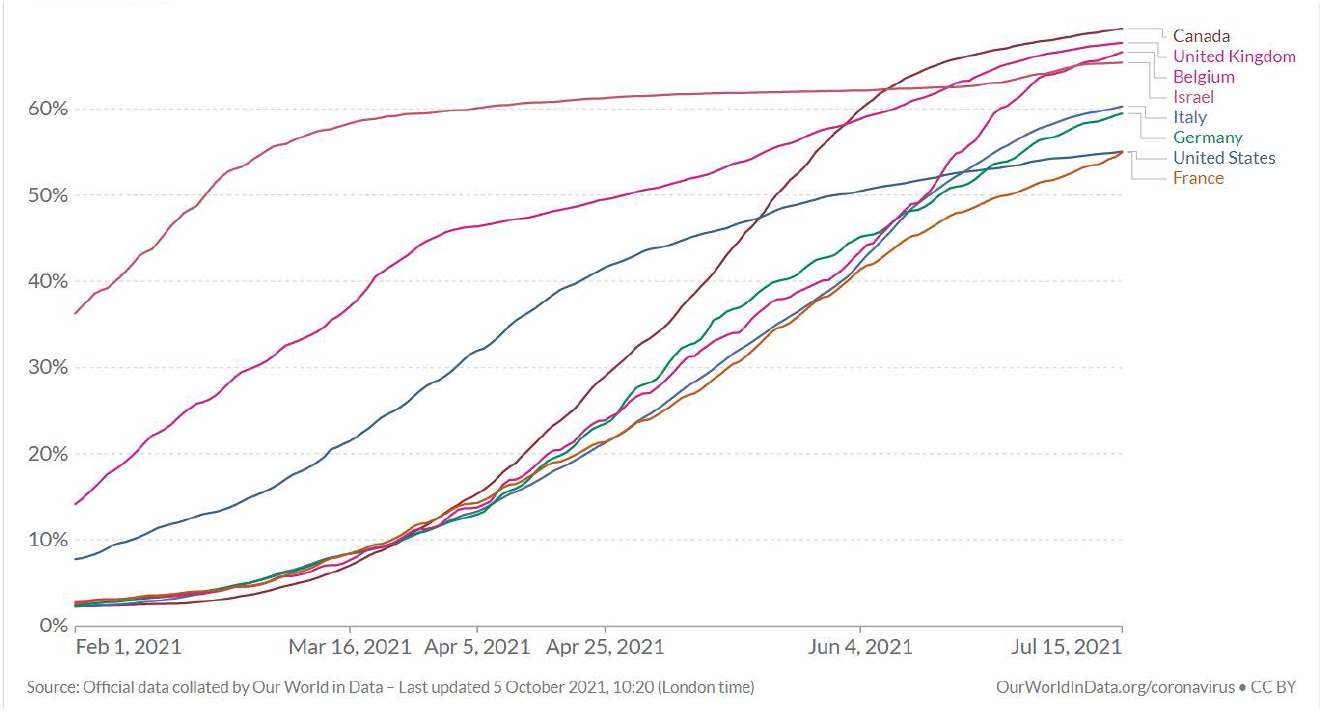
Share of people with at least one dose – example countries Notes: The figure plots the share of the population that has received at least one dose of a COVID-19 vaccine in selected countries. Source: [OurWorldinData](https://ourworldindata.org/).

**Extended Data Fig. 2.**
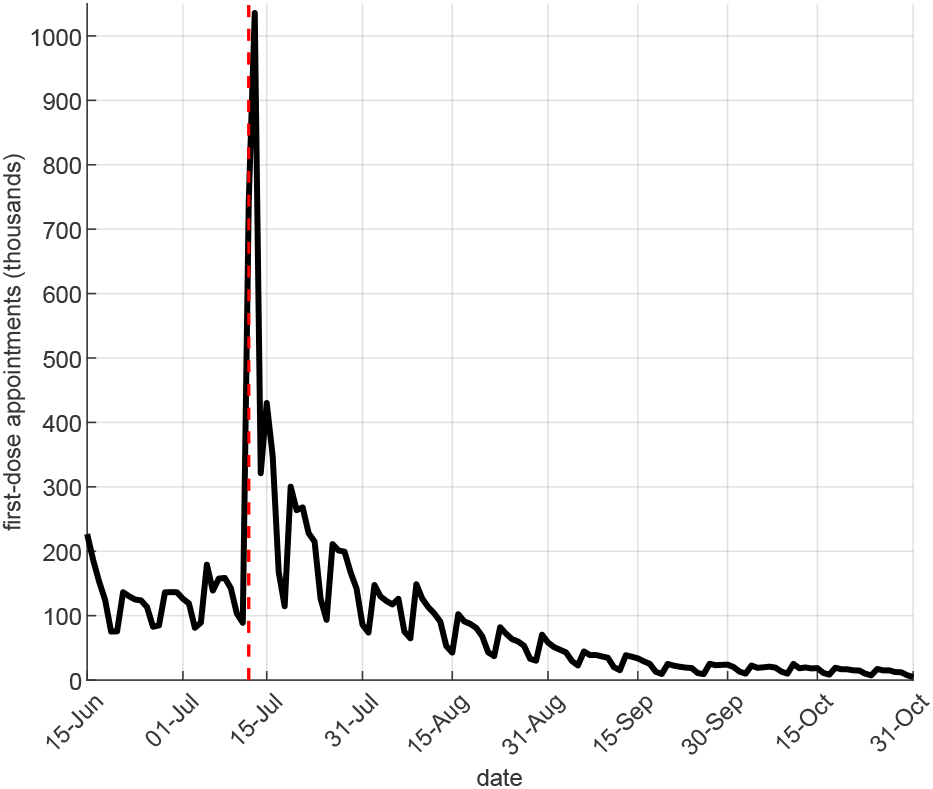
France – first-dose vaccination appointments Notes: The figure plots the daily first-dose vaccination appointments by date made on Doctolib, a booking website accounting for about 2/3 of cumulative COVID-19 vaccinations in France as of Q4 2021. Source: doctolib.fr. The vertical dashed line denotes the mandate announcement date, July 12, 2021.

**Extended Data Fig. 3.**
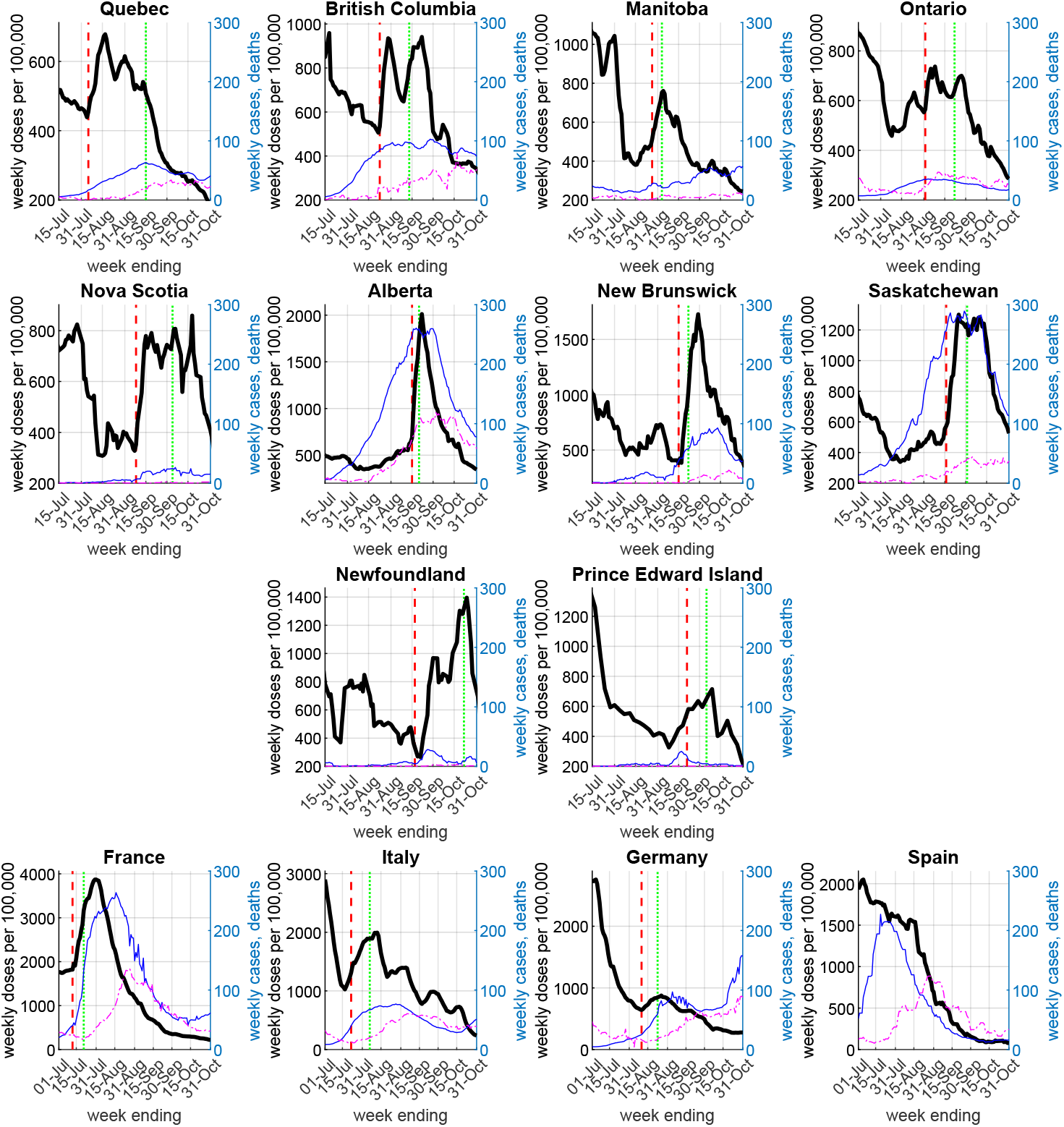
First doses per 100,000 people Notes: The figure plots the weekly administered COVID-19 vaccine first doses per 100,000 people for dates *t*− 6 to *t* (thick solid line), where *t* is the date on the horizontal axis. The vertical dashed lines denote the mandate announcement date for each province. The vertical dotted lines denote the mandate implementation (enforcement) date for each province (see Supplementary Table 1). The thin solid lines plot weekly cases per 100,000 (right axis) in each location; the thin dash-dotted lines plot weekly deaths (for the provinces) or weekly-averaged daily deaths (for the countries) (right axis).

**Extended Data Fig. 4.**
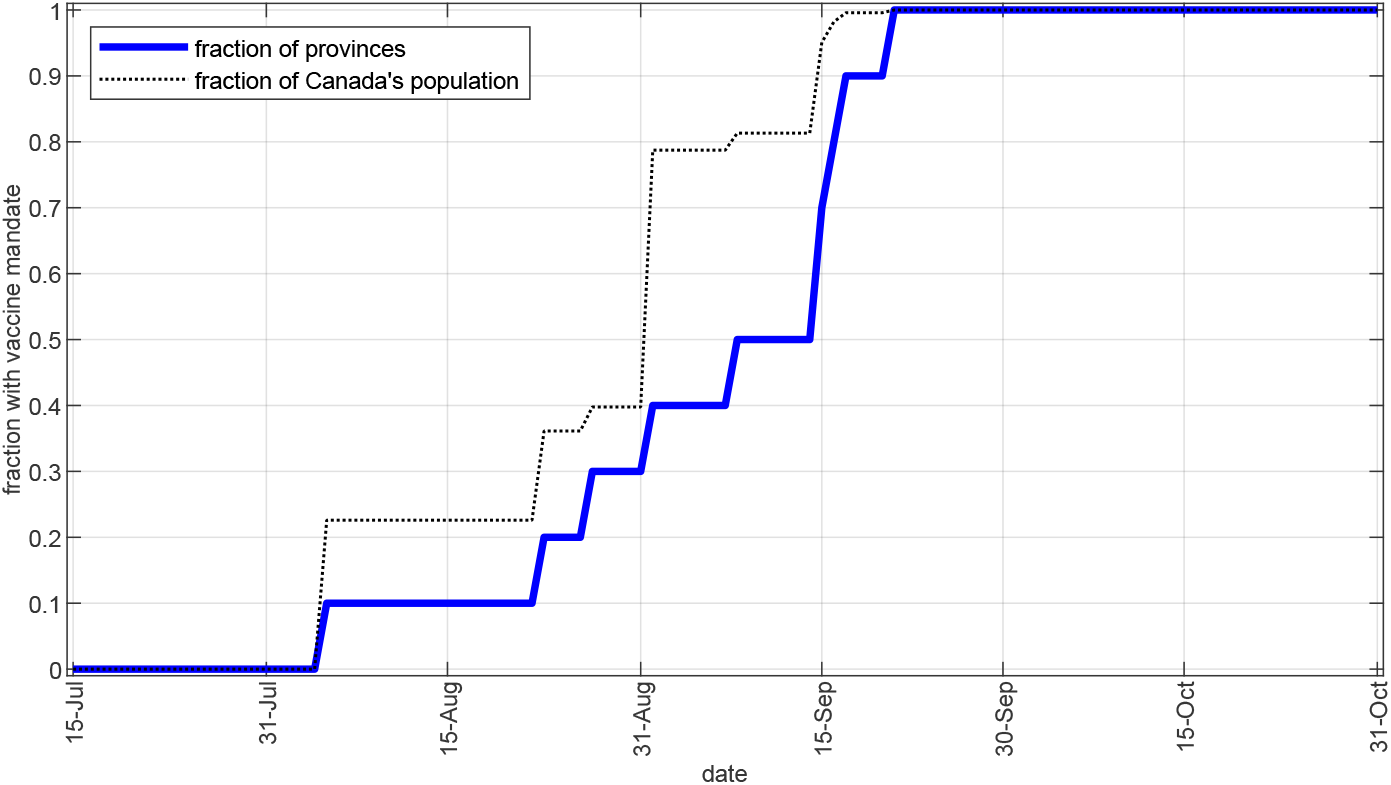
Canada – proof of vaccination mandates over time Notes: The figure plots the cumulative fraction of provinces and the cumulative fraction of Canada’s population for which a proof of vaccination mandate has been announced. See Supplementary Table 1 for the exact dates of mandate announcement in each province.

**Extended Data Fig. 5.**
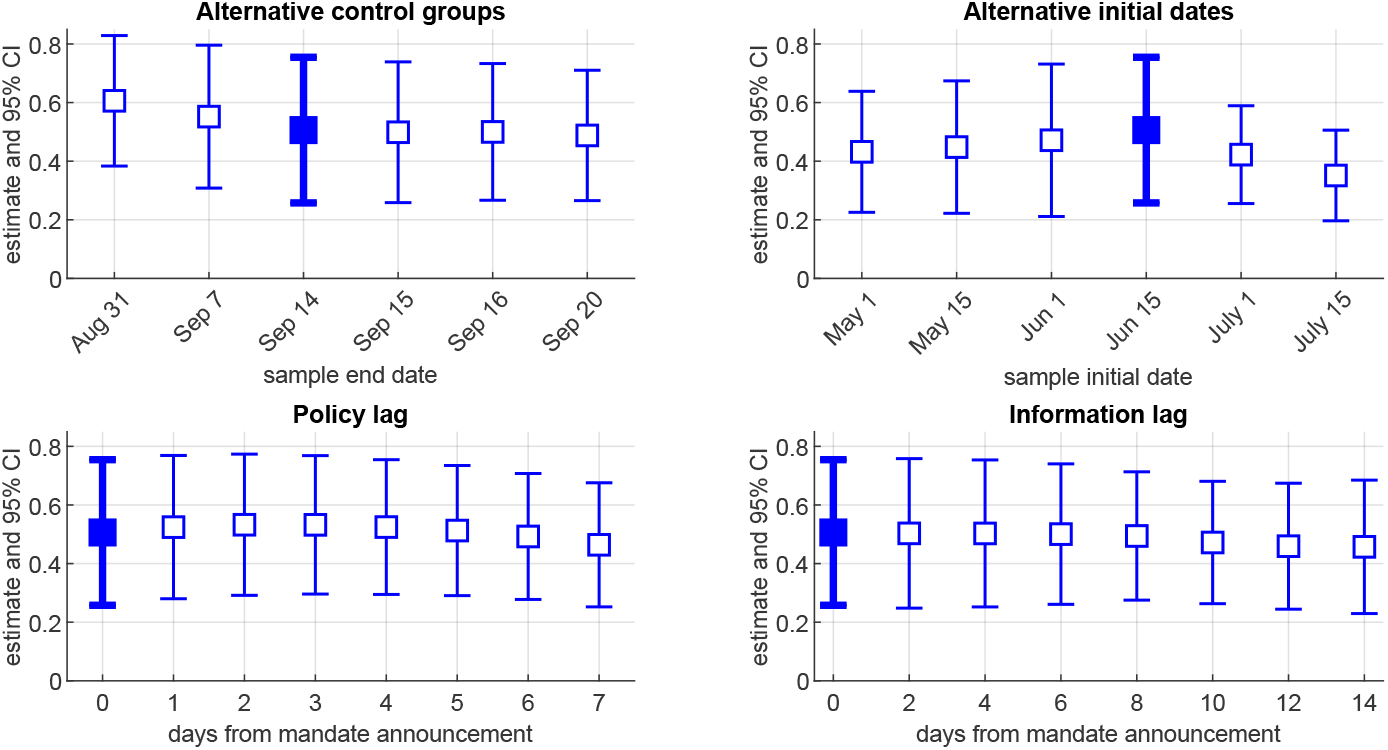
Robustness Notes: The figure plots the coefficient estimate, 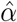 (denoted by square) and 95% confidence intervals (denoted by vertical error bars) of the mandate announcement variable *P*_*it*_ in equation (1). The upper left panel shows the estimates for different sample end dates and corresponding control groups including the baseline (Sep. 14, in bold). The upper right panel shows the estimates for different initial sample dates (May 1 to July 15, 2021), including the baseline (June 15, 2021, in bold). The lower left panel shows the estimates from a variant of equation (1) when using no lag in the policy announcement *P*_*it*_ (in bold, our baseline) and using lag of up to 7 days, i.e., *P*_*it*_−_*k*_ for *k* = 1, 2, *…,* 7. The lower right panel displays the estimates from a variant of equation (1) when using no lag for the information (cases and deaths) *I*_*it*_ (in bold, our baseline) or using lag of up to 14 days, i.e., *I*_*it*_−_*k*_ for *k* = 2, *…,* 14.

**Extended Data Fig. 6.**
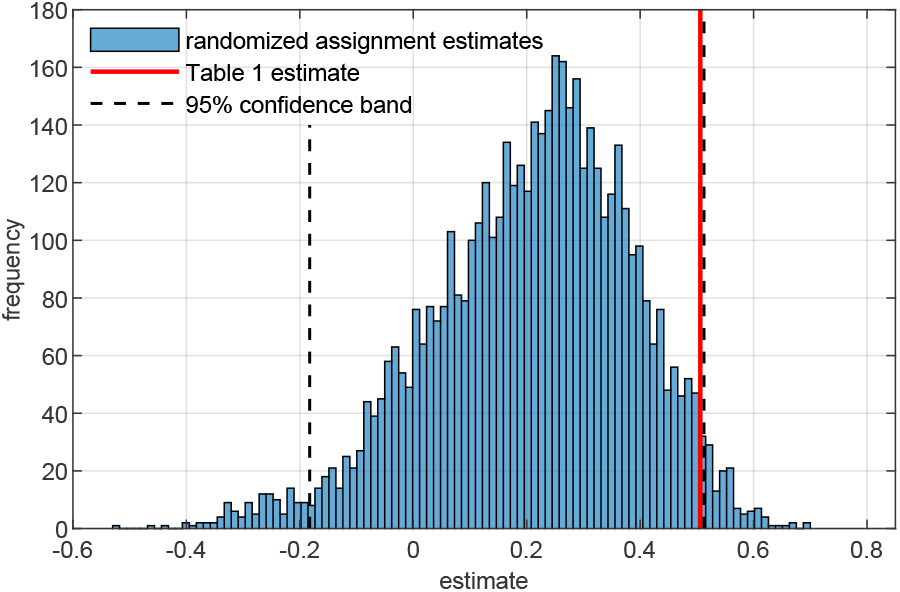
Random assignment of announcement dates Notes: We estimate equation (1) 5,000 times using the Sun and Abraham [23] treatment effect heterogeneity robust estimator after randomly assigning the date of mandate announcement for each province which has announced a mandate by Sep. 14. The figure plots the histogram of these placebo inference estimates, along with the 2.5-th and 97.5-th percentiles (dashed lines). The solid vertical line corresponds to the baseline estimate from column (2) in Table 1.

**Extended Data Fig. 7.**
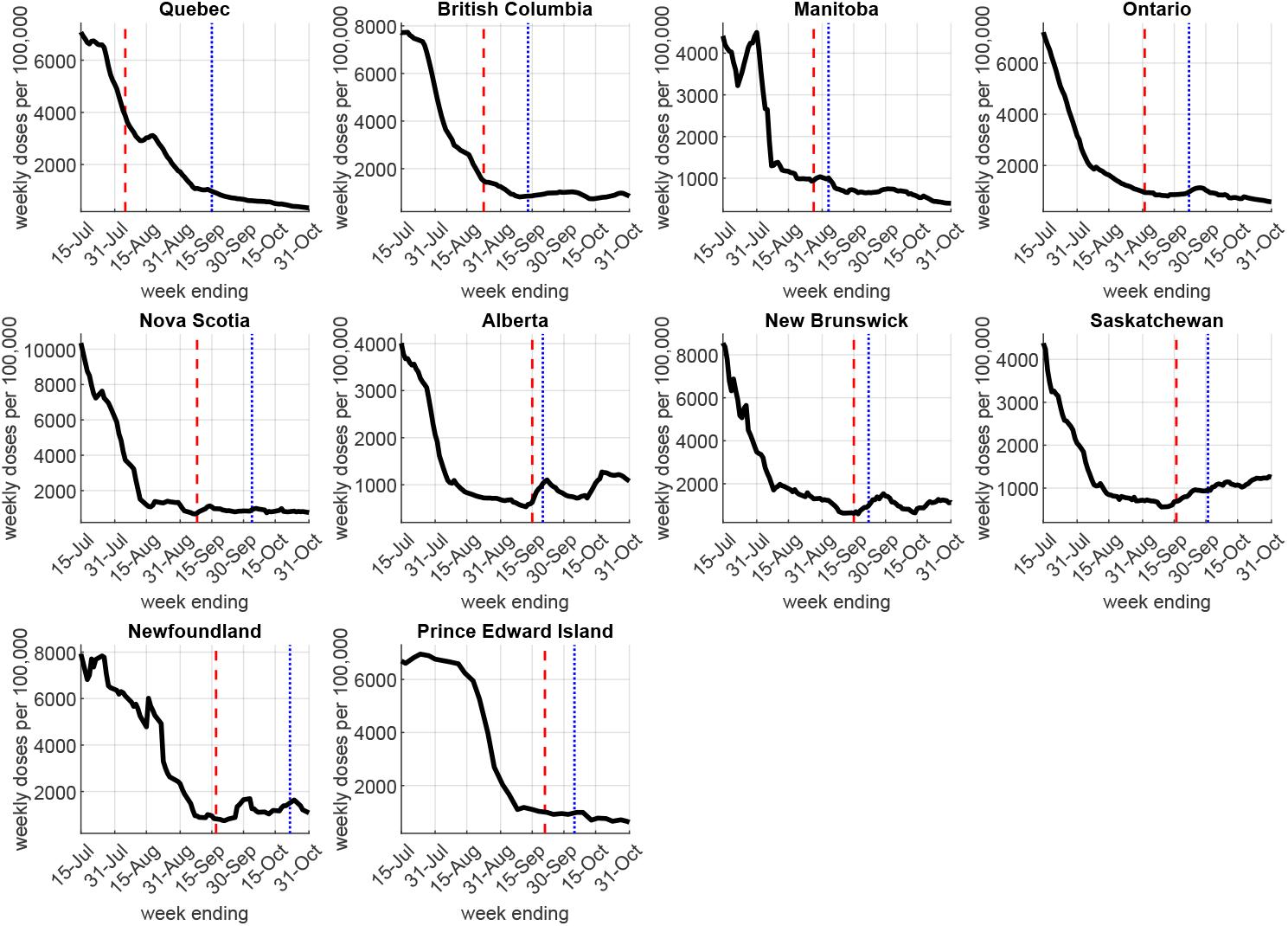
Canadian provinces - second doses per 100,000 people Notes: The figure plots the weekly administered COVID-19 vaccine *second* doses per 100,000 people for dates *t* − 6 to *t*, where *t* is the date on the horizontal axis. The vertical dashed lines denote the vaccination proof mandate announcement date for each province. The vertical dotted lines denote the mandate implementation (enforcement) date for each province (see Supplementary Table 1).

**Extended Data Fig. 8.**
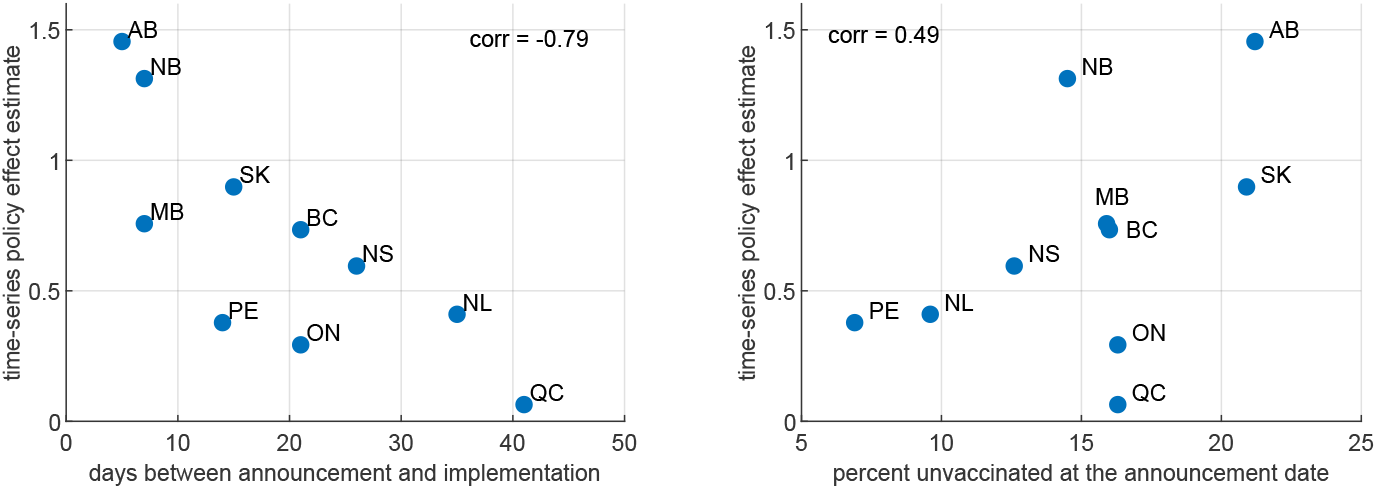
Time-series policy estimates – correlations Notes: The figure plots the time-series policy estimates, 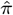 from Table 2 column (1) against the number of days between mandate announcement and implementation (left) and the percent remaining unvaccinated eligible people at the announcement date (right). The figure is for illustration; no causal claims are made.

**Extended Data Fig. 9.**
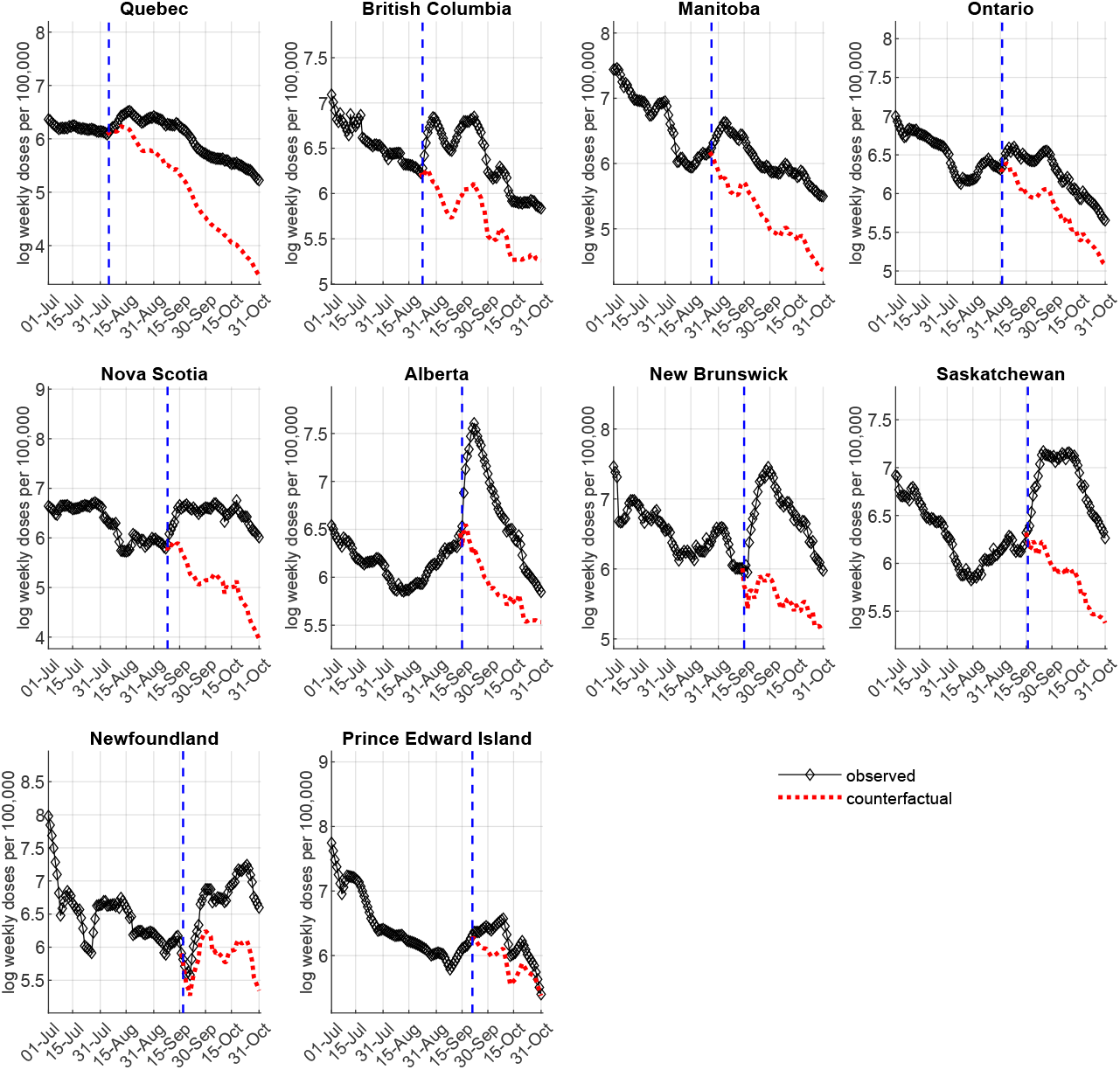
Canadian provinces – observed vs. no-mandate counterfactual weekly first doses as of October 31, 2021 (time-series estimates) Notes: The figure plots the observed (diamonds) and the estimated mean no-mandate counterfactual (dotted line) log weekly first doses per 100,000 people. We use the estimates from Table 2 to compute the counterfactuals, as specified in (3). The vertical dashed lines denote the mandate announcement date for each province.

**Extended Data Fig. 10.**
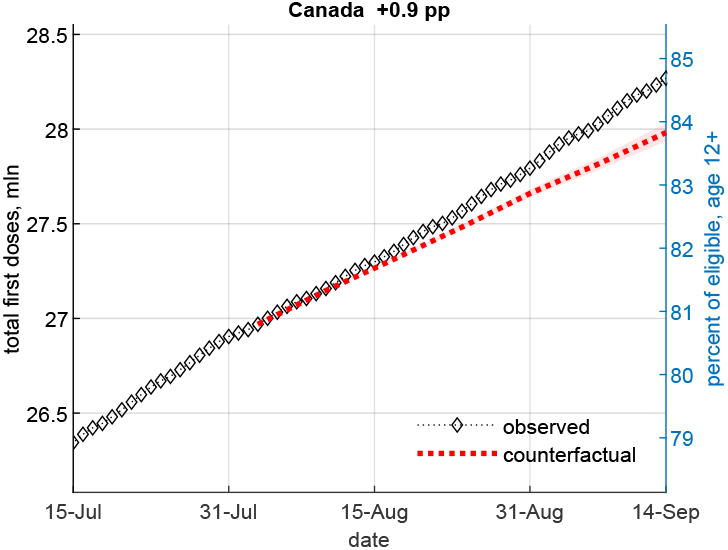
Canada – observed vs. no-mandate counterfactual first doses as of September 14, 2021 (DID estimate) Notes: The figure plots the observed (diamonds) and the estimated mean no-mandate counterfactual (dotted line) cumulative first doses (in millions) by date, with 5–95 percentile confidence bands (the shaded area). The counterfactual uses the *P*_*it*_ coefficient estimate from column (2) of Table 1. The number in the caption indicates the percentage point increase in first doses relative to the no-mandate counterfactual as of September 14, 2021.

## Supplementary information

### Robustness analysis details

#### Alternative initial dates

In Fig. 5, upper right panel, we show that our estimates of the effect of a mandate announcement on first dose vaccine uptake are not very sensitive to the choice of initial sample date between May 1 and Jul. 15, 2021. This provides further reassurance that vaccine supply constraints are not a major concern for our results.

#### Lags

In (1), we assume no lag between the mandate announcement *P*_*it*_ and a person’s ability to receive first dose (the outcome *V*_*it*_). In practice, a small delay may occur (e.g., from booking an appointment to receiving the vaccine) even in absence of supply constraints. The lower left panel of Fig. 5 displays the estimates 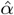 of the mandate effect when assuming zero lag (our baseline) to 7-day lag. The policy estimates remain large and statistically significant when varying the lag length, with a slight decrease for longer lags. The lower right panel of Fig. 5 displays the policy estimates 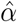 when assuming a lag of zero (our baseline) to 14 days in the information variables *I*_*it*_ (log weekly cases and deaths). The mandate effect estimates remains large and statistically significant when varying the lag length. We also checked a specification in which we lag the information variables *I*_*it*_ (log weekly cases and deaths) with the same lag as the policy *P*_*it*_, and the results are nearly identical.

#### Other robustness checks

We perform additional robustness checks in Table 5. For easier comparison, column (1) replicates the baseline estimates from columns (2) and (3) of Table 1. Column (2) of Table 5 omits cases and deaths, the coefficients for which are not significant in the baseline specification. In column (3), we control for log weekly deaths and log average weekly hospitalizations as information. Column (4) reports the estimates of a weighted specification using the provincial populations as weights and the baseline information variables (cases and deaths). The estimates are smaller than in the unweighted baseline (column 1), suggesting that smaller provinces contribute more to the the estimated effect of mandate announcements on vaccine uptake. In column (5), we re-estimate equation (1) using the standard OLS two-way fixed effects (TWFE) estimator which, as discussed in Methods, can be biased in the presence of heterogeneous treatment effects over cohorts and/or time. In our setting, we find that these estimates differ relatively little from our baseline Sun-Abraham treatment effect heterogeneity robust estimates.

In Table 5 column (6), we use log of daily (instead of weekly) first doses as the outcome and log of daily cases and deaths as information and find similar estimates to those in our baseline specification, although larger in week 0 since the outcome is not a moving sum. In column (7), we use the level of weekly first doses per 100,000 people as the outcome. The estimates are noisier than in the baseline and indicate that a mandate announcement increases weekly first dose uptake by roughly 482 per 100,000 people on average (CI -11–975, *p <* 0.06) after the announcement. Overall, all these alternative specifications confirm the robustness of our main finding of statistically significant and large impact of mandate announcements on vaccine uptake.

We also ran a specification replacing the province fixed effects with time-invariant province characteristics (GDP per capita, % rural population, population density, unemployment rate, ICU beds per capita, % of population of age 65 or older, full-time university students per capita, and political alignment). The estimated effect of the mandate announcements on vaccine uptake in this specification is very similar in magnitude and significance to our main estimates in Table 1.

#### Randomization inference

Difference-in-differences inference from the model in equation (1) relies on asymptotic approximations requiring a sufficiently large number of provinces. While we do account for the fact that there are only 10 provinces in our sample by using wild bootstrap to compute the estimates’ p-values, as an additional robustness check, we implement a variant of Fisher’s randomization test, [44]. Specifically, we estimate equation (1) 5,000 times by randomly assigning the mandate announcement date for each of the ‘treated’ provinces in our baseline specification. Fig. 6 plots the histogram of the randomized inference (placebo) mandate effect estimates, along with the 95% confidence band. Only 2.88% of the 5,000 placebo estimates are larger than our baseline estimate 0.506 from column (2) of Table 1 (the solid vertical line on Fig. 6), offering assurance that our main result does not hinge on asymptotic inference.

#### Mandate implementation

Our main results use the regional variation in the timing of announcement of proof of vaccination mandates to examine their effect on vaccine uptake. In addition, there is variation in the interval between the mandates’ announcement and implementation (coming into force), as well as in the implementation dates themselves – see Table 1 and Fig. 3. We checked whether we can detect additional effects on first-dose uptake, associated with the mandates’ implementation, by adding to equation (1) an indicator variable that equals to 1 post-implementation and 0 otherwise. The mandates were implemented in all provinces between Sep. 3 and Oct. 22, 2021. The estimate on this indicator variable is positive but not statistically significantly different from zero (not displayed). However, we cannot conclude that there do not exist implementation effects, since even the latest feasible sample end date (Sep. 20, 2021) necessitated by the requirement to have a not yet treated control group is only soon after the mandate implementation date in 3 of the 10 provinces (see Table 1).

#### Potential spillover effects

To check for potential spillover effects whereby a mandate in one province may have an effect on vaccine uptake in another province, we ran the time-series regression, equation (2), for Ontario and British Columbia (BC) assuming earlier announcement dates from other provinces. Specifically, the regression for Ontario using BC’s announcement date (Aug. 23) instead of Ontario’s actual announcement date (Sep. 1) yields a statistically insignificantly different from zero policy estimate, 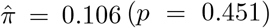. Similarly, regressions for BC and Ontario using the Quebec announcement date (Aug. 5) yield 0.182 (*p* = 0.263) and -0.451 (*p* = 0.019) respectively. While this evidence does not support the presence of spillover effects, if they did exist across some provinces or dates, then we can interpret our estimates as a lower bound of the full policy effect, as some fraction of people would have been vaccinated already before the announcements.

#### Time-series fit and out-of-sample projection

In Supplementary Fig. 1, we display the time-series model fit with the data until Oct. 31, 2021. In addition, we plot out-of-sample predictions obtained using equation (2), the estimates from Table 2 and the actual weekly cases and deaths data for these out-of-sample dates. The time-series specification fits the weekly first dose data very well (e.g., the adjusted R-squared is 0.997 for France, 0.940 for Italy and 0.990 for Germany), and the out-of-sample projections (dotted line) remain close to the observed first-dose data from November, with the exception of Germany, where the COVID-19 certificate became more restrictive in this period by excluding unvaccinated people that were previously able to supply a negative test from indoor dining, bars, hairdressers, etc. A constraining factor for the out-of-sample projections is that first doses for the 5-to-11 age group were approved in November (Nov. 19 in Canada and Nov. 25 in the EU), and, for many jurisdictions in our sample, we cannot distinguish these doses from adult first doses in the vaccinations data. Hence, we only show out-of-sample projections for three additional weeks beyond our end-sample date, until Nov. 21, 2021.

#### Time-series – placebo announcement dates

Similar to the DID randomization inference robustness exercise (Fig. 6), we use our time-series model to estimate the mandate policy effect (the parameter *π* in equation (2)) for a range of placebo announcement dates (132 unique dates in total including the true announcement date), from Jun. 17 to Oct. 24 for each of France, Italy and Germany. Supplementary Fig. 2 displays the results. There is only a small number of placebo dates (mostly in a close neighbourhood of the true announcement date) for which the time-series regression yields a larger policy estimate. Specifically, there are 12 dates (9.1%) with estimate larger than that for the true announcement date for France, 14 dates (10.6%) for Italy and only 5 such dates (3.8%) for Germany. While these results are reassuring (although relatively noisy for France and Italy), they should only be treated as supplementary, since the time-series approach and specification we use rely on a known date for the policy regime change.

#### Related literature – details

[26] use a synthetic control approach to evaluate the impact of COVID-19 certification mandates on vaccine uptake, overall and by age group, in six countries. The authors estimate significant increases in vaccinations in France (about 8.6 mln), Italy (4 mln) and Israel (2.1 mln) from 20 days prior to 40 days after the mandate implementation. While the main takeaway is similar, our paper differs in several important ways. First, we provide robust difference-in-differences evidence using the variation in mandate timing within the same country, Canada, in addition to evidence using time-series methods for France, Italy and Germany. Second, instead of total vaccinations, we focus on first doses, as most directly reflecting the decision to become immunized and avoiding potential issues related to second dose scheduling or availability in spring/summer 2021. This is one possible reason why, unlike [26], we find a statistically significant increase in vaccine uptake in Germany around the mandate. When using total vaccinations instead of first doses, we also do not find a statistically significant effect for Germany. Third, instead of using a fixed 20-day cutoff before implementation, we use the actual mandate announcement date (the interval between mandate announcement and implementation in the ten Canadian provinces and three countries in our sample varies from 5 to 41 days) as the intervention indicator and show a strong impact on vaccine uptake thereafter.

There is little evidence that financial or behavioural nudges increase the vaccination rate among hesitant people. [12] report results from a mid-2021 randomized controlled trial (RCT) with unvaccinated members of a large Medicare health plan in a California county with 77% vaccination rate at the time of the study. The authors examine the effect of $10 or $50 financial incentives, different public health messages and an appointment scheduler, on vaccination intentions and uptake within 30 days of the intervention. They find that none of the financial or behavioural treatments increased the vaccination rate among the treated. The proof-of-vaccination mandates we analyze target a similar group of unvaccinated people that have had the opportunity to be vaccinated for a long time. The population-weighted average first-dose vaccination rate on the dates of the provincial mandate announcements is 83.3%, even higher than in [12], suggesting that financial or behavioural incentives are even less likely to be effective in our setting. In an RCT study in Sweden, [13] find that a modest payment of SEK 200 (USD 24) is associated with a 4.2 p.p. increase in vaccinations, relative to a baseline rate of 71.6% in the control group, while none of three behavioural nudges had impact. However, the RCT subjects were yet to become eligible to be vaccinated, and hence the setting is not comparable to ours in which weeks have elapsed after widespread vaccine availability.

**Supplementary Fig. 1.**
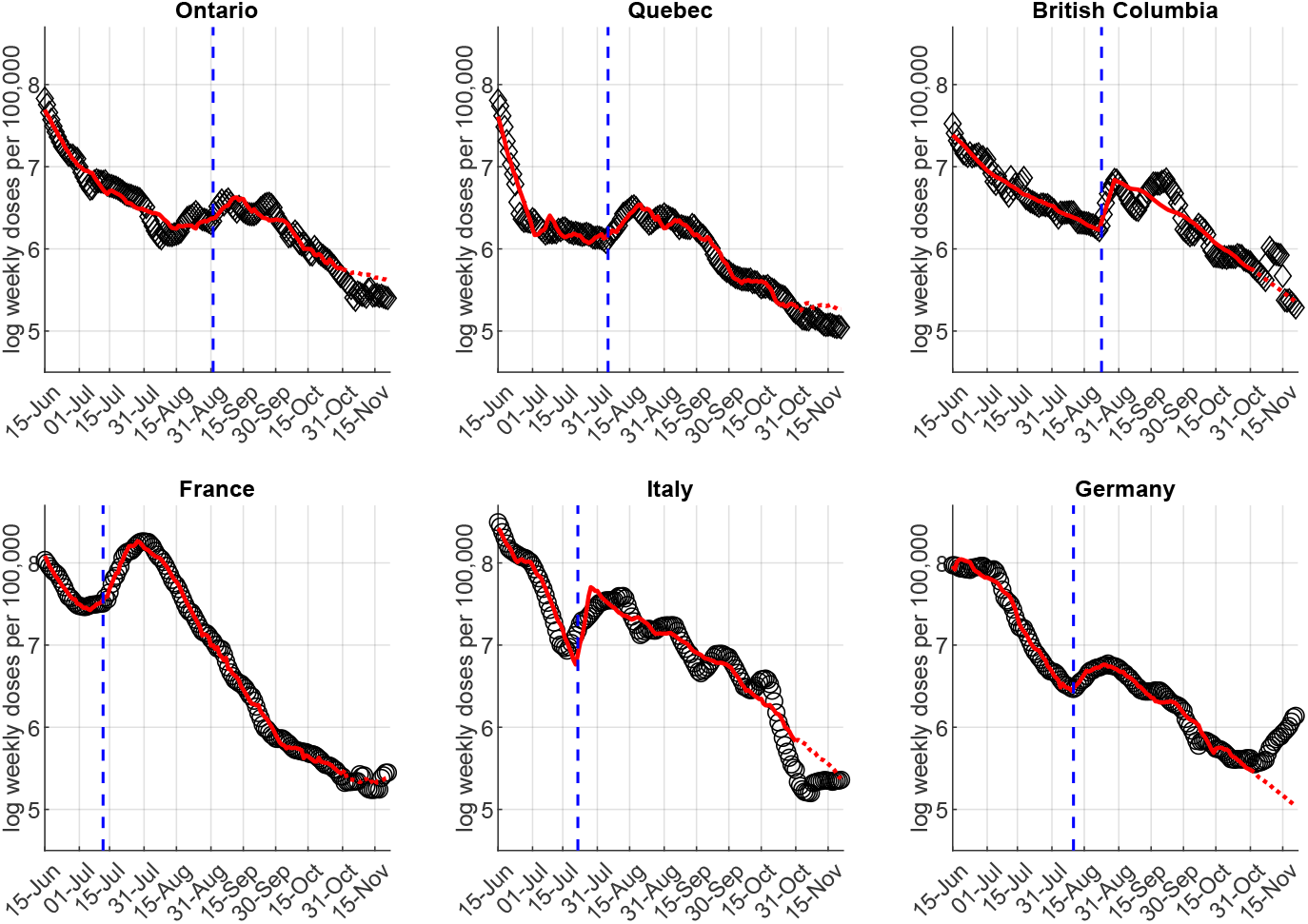
Time-series fit and out-of-sample projection Notes: The circles are log weekly first doses per 100,000 people. The solid line is the fitted value from the time-series regression, equation (2). The dotted line is the out-of-sample extrapolation of the fitted value line for three additional weeks, using cases and deaths data until Nov. 21, 2021. Germany introduced a stricter version of their COVID certificate in November 2021.

**Supplementary Fig. 2.**
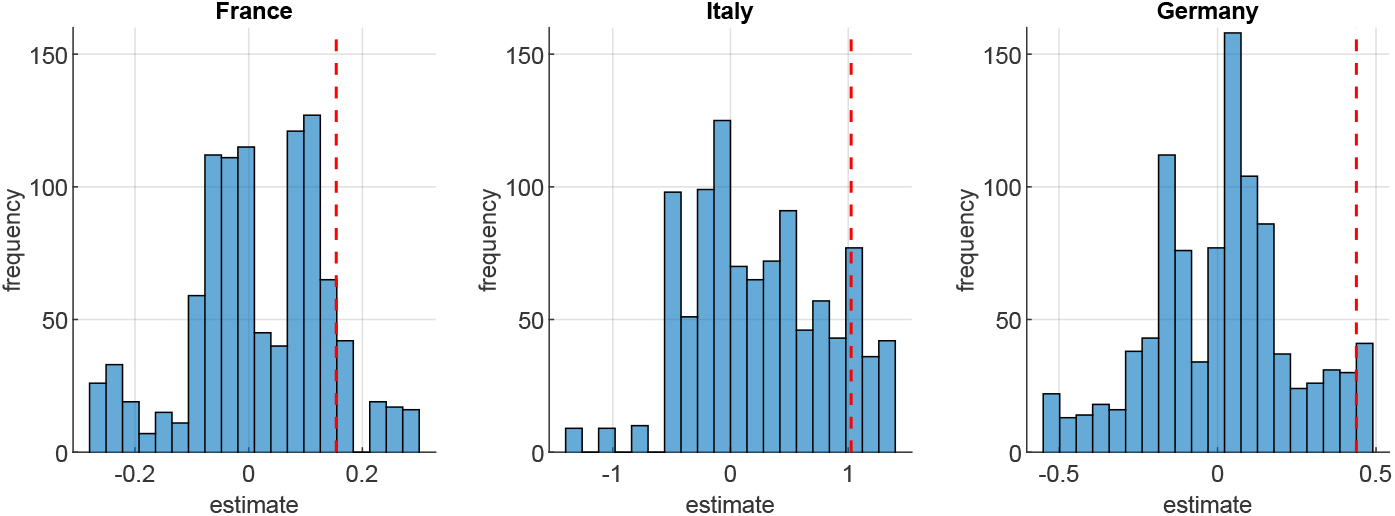
Time-series – placebo announcement dates Notes: The figure reports time-series estimates for the mandate policy effect (the parameter *π* in equation (2)) from 132 runs with all possible placebo mandate announcement dates between June 17 and Oct 24. The vertical dashed line is the main estimate from Table 2 using the actual mandate announcement date.

**Supplementary Table 1.**
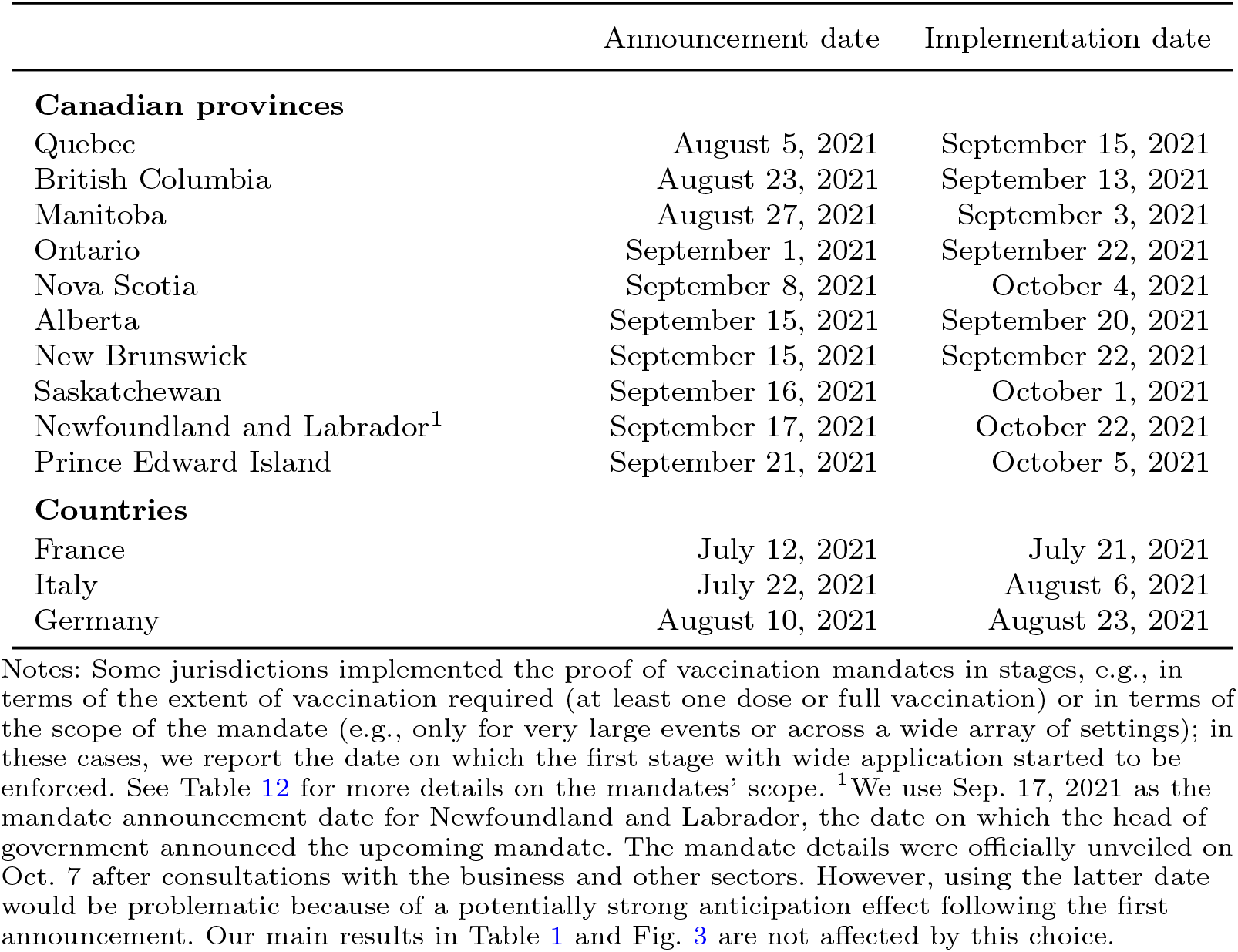
Mandate announcement and implementation dates

**Supplementary Table 2.** Eligible population (age 12 or older)

|  | Total eligible | Percent unvaccinated on<br>mandate announcement date |
| --- | --- | --- |
| Quebec | 7,532,499 | 16.3% |
| British Columbia | 4,644,795 | 16.0% |
| Manitoba | 1,176,020 | 15.9% |
| Ontario | 13,038,060 | 16.3% |
| Nova Scotia | 884,351 | 12.6% |
| Alberta | 3,791,765 | 21.2% |
| New Brunswick | 701,871 | 14.5% |
| Saskatchewan | 996,908 | 20.9% |
| Newfoundland and Labrador | 467,761 | 9.6% |
| Prince Edward Island | 145,432 | 6.9% |
| France | 57,668,019 | 37.0% |
| Italy | 54,036,939 | 31.2% |
| Germany | 74,100,000 | 29.3% |

Notes: The table reports the number of vaccine eligible people (age 12 or older) by province or country. Sources: Canada (link), France (link), Italy (link), Germany (link).

**Supplementary Table 3.** Canadian provinces – age 12-plus eligibility dates

|  |  |
| --- | --- |
| Quebec | May 25, 2021 |
| British Columbia | May 19, 2021 |
| Manitoba | May 14, 2021 |
| Ontario | May 23, 2021 |
| Nova Scotia | May 27, 2021 |
| Alberta | May 10, 2021 |
| New Brunswick | May 26, 2021 |
| Saskatchewan | May 20, 2021 |
| Newfoundland and Labrador | May 17, 2021 |
| Prince Edward Island | May 18, 2021 |

Notes: The table reports the first date at which any person of age 12 or higher was eligible to register and receive first dose of a COVID-19 vaccine in the respective province.

**Supplementary Table 4.** Canadian provinces - different control groups

| end date<br>control group<br>p-values in [ ] | Outcome: log weekly vaccine first doses, $V_{it}$ | | | | | |
| --- | --- | --- | --- | --- | --- | --- |
|  | Sep. 20<br>PE<br>(1) | Sep. 16<br>(1)+NL<br>(2) | Sep. 15<br>(2)+SK<br>(3) | Sep. 14<br>(3)+AB,NB<br>(4) | Sep. 7<br>(4)+NS<br>(5) | Aug. 31<br>(5)+ON<br>(6) |
| week 0 | 0.372 ***<br>[0.001] | 0.364 **<br>[0.019] | 0.351 **<br>[0.017] | 0.359 ***<br>[0.005] | 0.384 ***<br>[0.009] | 0.404 ***<br>[0.000] |
| week 1 | 0.561 ***<br>[0.001] | 0.552 ***<br>[0.001] | 0.545 ***<br>[0.001] | 0.543 ***<br>[0.001] | 0.620 ***<br>[0.002] | 0.837 ***<br>[0.000] |
| week 2 | 0.442 **<br>[0.029] | 0.469 **<br>[0.025] | 0.477 **<br>[0.019] | 0.498 ***<br>[0.010] | 0.687 ***<br>[0.000] | 0.767 ***<br>[0.000] |
| week 3 | 0.540 ***<br>[0.006] | 0.705 ***<br>[0.002] | 0.695 ***<br>[0.001] | 0.705 ***<br>[0.001] | 0.739 ***<br>[0.002] | 0.712 ***<br>[0.002] |
| week 4 | 0.690 **<br>[0.025] | 0.712 **<br>[0.018] | 0.712 **<br>[0.018] | 0.713 ***<br>[0.018] | 0.710 **<br>[0.026] |  |
| week 5 | 0.650 *<br>[0.059] | 0.651 *<br>[0.059] | 0.642 *<br>[0.060] | 0.651 *<br>[0.056] |  |  |
| week 6+ | 0.492 *<br>[0.089] | 0.651 *<br>[0.060] |  |  |  |  |
| R-squared | 0.811 | 0.818 | 0.820 | 0.821 | 0.829 | 0.839 |
| sample size, N | 980 | 940 | 930 | 920 | 850 | 780 |
| province fixed effects | X | X | X | X | X | X |
| date fixed effects | X | X | X | X | X | X |
| cases and deaths | X | X | X | X | X | X |

Notes: Sun and Abraham, [23], treatment effect heterogeneity robust estimates. The table reports estimates for different treatment and control (latest treated) groups of provinces which determine the sample end date. “week n”, where $n = 0, 1, 2, \dots$ , is a binary variable that takes value 1 for the days in the n-th week immediately after the announcement date (week 0 is the week starting at the announcement date) and value 0 otherwise. The cases and deaths variables are log weekly totals for dates $t - 6$ to $t$ . P-values from wild bootstrap (boottest) standard errors clustered by province with 4,999 repetitions are reported in the square brackets. \*, \*\* and \*\*\* denote 10%, 5% and 1% significance (two-sided) respectively.

**Supplementary Table 5.** Robustness

| p-values in [ ] | Outcome: log weekly first vaccine doses, $V_{it}$ | | | | | | |
| --- | --- | --- | --- | --- | --- | --- | --- |
|  | baseline | no cases<br>& deaths | deaths &<br>hospitaliz. | population<br>weighted | OLS<br>TWFE | daily<br>outcome | level<br>outcome |
|  | (1) | (2) | (3) | (4) | (5) | (6) | (7) |
| **A. All post-announcement dates** |  |  |  |  |  |  |  |
| mandate announced $P_{it}$ | 0.506 ***<br>[0.001] | 0.504 ***<br>[0.002] | 0.507 ***<br>[0.000] | 0.363 ***<br>[0.003] | 0.494 **<br>[0.020] | 0.513 ***<br>[0.001] | 481.6 *<br>[0.052] |
| R-squared | 0.820 | 0.817 | 0.821 | 0.888 | 0.812 | 0.577 | 0.662 |
| **B. By week after the mandate announcement** |  |  |  |  |  |  |  |
| week 0 | 0.359 ***<br>[0.005] | 0.344 **<br>[0.020] | 0.362 ***<br>[0.002] | 0.229 **<br>[0.027] | 0.378 ***<br>[0.007] | 0.517 ***<br>[0.001] | 309.9 *<br>[0.082] |
| week 1 | 0.543 ***<br>[0.001] | 0.546 ***<br>[0.003] | 0.545 ***<br>[0.000] | 0.328 *<br>[0.088] | 0.570 **<br>[0.018] | 0.394 ***<br>[0.002] | 505.3 *<br>[0.056] |
| week 2 | 0.498 **<br>[0.010] | 0.509 **<br>[0.016] | 0.503 **<br>[0.010] | 0.335 *<br>[0.056] | 0.520 *<br>[0.054] | 0.431 ***<br>[0.000] | 510.0 *<br>[0.088] |
| week 3 | 0.705 ***<br>[0.001] | 0.693 ***<br>[0.001] | 0.693 ***<br>[0.000] | 0.451 ***<br>[0.000] | 0.677 ***<br>[0.004] | 0.672 ***<br>[0.001] | 676.3 *<br>[0.027] |
| week 4 | 0.713 **<br>[0.018] | 0.723 **<br>[0.020] | 0.705 ***<br>[0.008] | 0.370<br>[0.109] | 0.672 **<br>[0.022] | 0.758 ***<br>[0.003] | 754.1 *<br>[0.068] |
| weeks 5+ | 0.651 *<br>[0.056] | 0.678 **<br>[0.041] | 0.658 **<br>[0.046] | 0.286<br>[0.423] | 0.650 *<br>[0.052] | 0.510 *<br>[0.060] | 732.3<br>[0.119] |
| R-squared | 0.821 | 0.819 | 0.823 | 0.896 | 0.816 | 0.580 | 0.663 |
| sample size, N | 920 | 920 | 920 | 920 | 920 | 920 | 920 |
| province fixed effects | X | X | X | X | X | X | X |
| date fixed effects | X | X | X | X | X | X | X |

Notes: Sun and Abraham, [23], treatment effect heterogeneity robust estimates. Sample period: June 15 to September 14, 2021 using Alberta, New Brunswick, Saskatchewan, Prince Edward Island and Newfoundland as control group (latest treated); “week $n$ ”, where $n = 0, 1, 2, \dots$ , is a binary variable that takes value 1 for the days in the $n$ -th week immediately after the announcement date (week 0 is the week starting at the announcement date) and value 0 otherwise. All specifications include log weekly cases $C_{it}$ and log weekly deaths $D_{it}$ as information controls unless otherwise specified. P-values from wild bootstrap (boottest) standard errors clustered by province with 4,999 repetitions are reported in the square brackets. Column (1) reproduces the baseline estimates from columns (2)–(3) of Table 1. Column (2) does not control for cases and deaths. Column (3) uses log average weekly hospitalizations and log weekly deaths as information. Column (4) reports the estimates from a weighted version of equation (1) with the provinces’ populations used as weights. Column (5) displays results from estimating (1) via the standard OLS two-way fixed effects (TWFE) method. Column (6) uses log *daily* first doses as the outcome. Column (7) uses the *level* of weekly first doses per 100,000 people as the outcome. \*, \*\* and \*\*\* denote 10%, 5% and 1% significance (two-sided) respectively.

**Supplementary Table 6.** Robustness – standard errors

| Outcome: log weekly first doses, $V_{it}$ | | | |
| --- | --- | --- | --- |
|  | (1) | (2) |  |
| mandate announced | 0.506<br>(0.001) ***<br>[0.001] ***<br>{0.000} *** |  |  |
| week 0 |  | 0.359<br>(0.004) ***<br>[0.005] ***<br>{0.005} *** |  |
| week 1 |  | 0.543<br>(0.002) ***<br>[0.001] ***<br>{0.000} *** |  |
| week 2 |  | 0.498<br>(0.006) ***<br>[0.010] **<br>{0.009} *** |  |
| week 3 |  | 0.705<br>(0.000) ***<br>[0.001] ***<br>{0.001} *** |  |
| week 4 |  | 0.713<br>(0.003) ***<br>[0.018] **<br>{0.016} ** |  |
| week 5+ |  | 0.651<br>(0.011) **<br>[0.056] *<br>{0.055} * |  |
| R-squared | 0.820 | 0.821 |  |
| sample size, N | 920 | 920 |  |
| province fixed effects | X | X |  |
| date fixed effects | X | X |  |
| cases and deaths | X | X |  |

Notes: P-values from standard clustering (Stata command “cluster”) by province in the ( ) parentheses; wild bootstrap (Stata command “boottest”) with one-way clustering by province and 4,999 repetitions in the [ ] square brackets, and wild bootstrap with two-way clustering by province and date with 4,999 repetitions in the { } curly braces. “week $n$ ”, where $n = 0, 1, 2, \dots$ , is a binary variable that takes value 1 for the days in the $n$ -th week immediately after the announcement date (week 0 is the week starting at the announcement date) and value 0 otherwise. The cases and deaths variables are log weekly totals for dates $t - 6$ to $t$ . \*, \*\* and \*\*\* denote 10%, 5% and 1% significance (two-sided) respectively.

**Supplementary Table 7.** Second doses

| Outcome: log weekly vaccine second doses, $S_{it}$ | | | |
| --- | --- | --- | --- |
| p-values in [ ] | (1) | (2) | (3) |
| mandate announced, $P_{it}$ | 0.099 | 0.116 | |
|  | [0.405] | [0.298] |  |
| week 0 |  |  | 0.041 |
|  |  |  | [0.630] |
| week 1 |  |  | 0.117 |
|  |  |  | [0.252] |
| week 2 |  |  | 0.145 |
|  |  |  | [0.195] |
| week 3 |  |  | 0.201 |
|  |  |  | [0.176] |
| week 4 |  |  | 0.177 |
|  |  |  | [0.243] |
| week 5+ |  |  | 0.185 |
|  |  |  | [0.120] |
| R-squared | 0.848 | 0.853 | 0.854 |
| sample size, N | 920 | 920 | 920 |
| province fixed effects | X | X | X |
| date fixed effects | X | X | X |
| log weekly cases and deaths |  | X | X |

Notes: Sun and Abraham, [23], treatment effect heterogeneity robust estimates. Time period: June 15 to September 14, 2021 using Alberta, New Brunswick, Saskatchewan, Prince Edward Island and Newfoundland as control group (latest treated); “week n”, where $n = 0, 1, 2, \dots$ , is a binary variable that takes value 1 for the days in the n-th week immediately after the announcement date (week 0 is the week starting at the announcement date) and value 0 otherwise. The cases and deaths variables are log weekly totals for dates $t - 6$ to $t$ . P-values from wild bootstrap (boottest) standard errors clustered by province with 5,000 repetitions are reported in the square brackets. \*, \*\* and \*\*\* denote 10%, 5% and 1% significance (two-sided) respectively.

**Supplementary Table 8.**
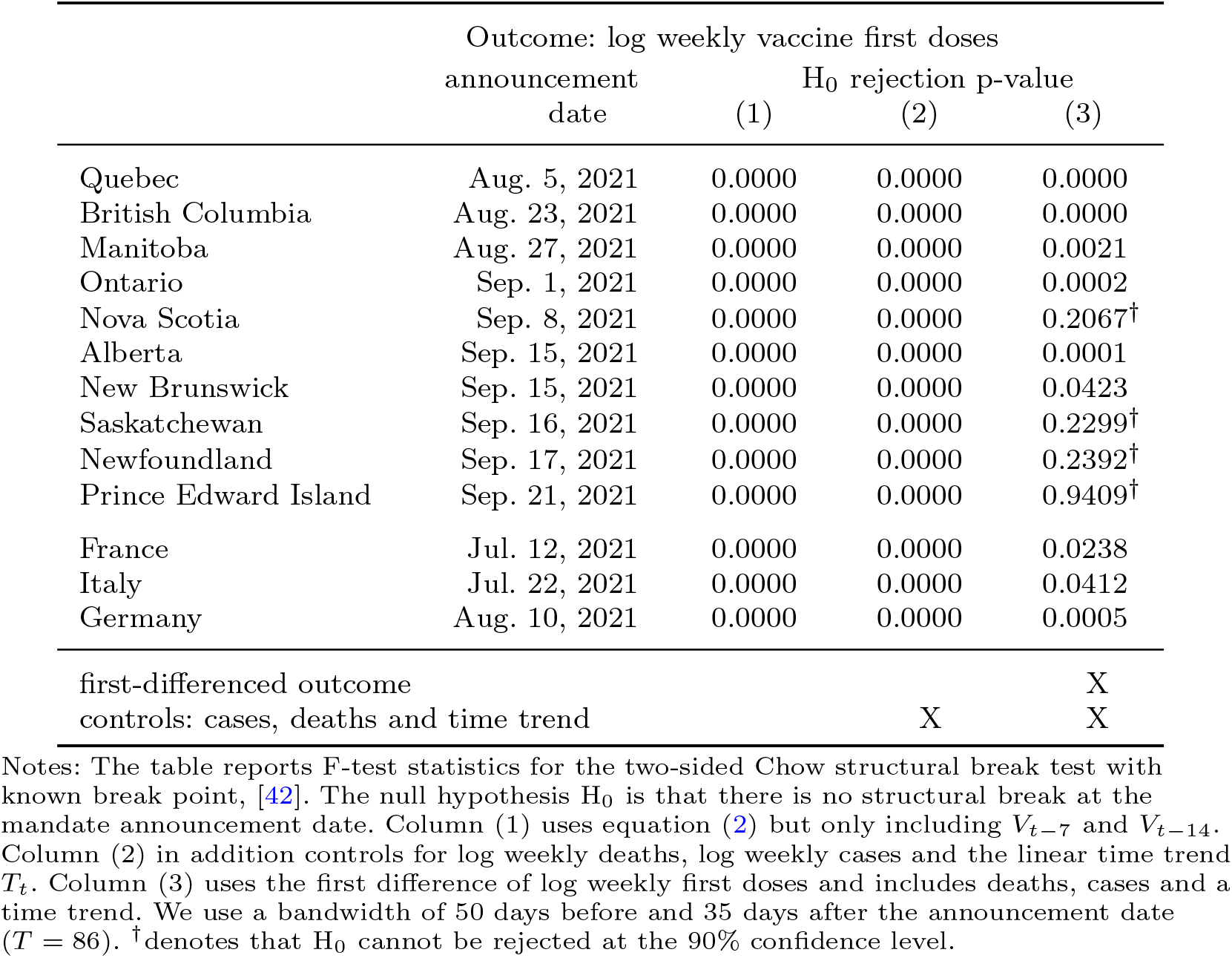
Structural break at the mandate announcement date

|  | Outcome: log weekly vaccine first doses<br>announcement date | H<sub>0</sub> rejection p-value |  |  |
| --- | --- | --- | --- | --- |
|  |  | (1) | (2) | (3) |
| Quebec | Aug. 5, 2021 | 0.0000 | 0.0000 | 0.0000 |
| British Columbia | Aug. 23, 2021 | 0.0000 | 0.0000 | 0.0000 |
| Manitoba | Aug. 27, 2021 | 0.0000 | 0.0000 | 0.0021 |
| Ontario | Sep. 1, 2021 | 0.0000 | 0.0000 | 0.0002 |
| Nova Scotia | Sep. 8, 2021 | 0.0000 | 0.0000 | 0.2067<sup>†</sup> |
| Alberta | Sep. 15, 2021 | 0.0000 | 0.0000 | 0.0001 |
| New Brunswick | Sep. 15, 2021 | 0.0000 | 0.0000 | 0.0423 |
| Saskatchewan | Sep. 16, 2021 | 0.0000 | 0.0000 | 0.2299<sup>†</sup> |
| Newfoundland | Sep. 17, 2021 | 0.0000 | 0.0000 | 0.2392<sup>†</sup> |
| Prince Edward Island | Sep. 21, 2021 | 0.0000 | 0.0000 | 0.9409<sup>†</sup> |
| France | Jul. 12, 2021 | 0.0000 | 0.0000 | 0.0238 |
| Italy | Jul. 22, 2021 | 0.0000 | 0.0000 | 0.0412 |
| Germany | Aug. 10, 2021 | 0.0000 | 0.0000 | 0.0005 |
| first-differenced outcome |  |  |  | X |
| controls: cases, deaths and time trend |  |  | X | X |

Notes: The table reports F-test statistics for the two-sided Chow structural break test with known break point, [42]. The null hypothesis $H_0$ is that there is no structural break at the mandate announcement date. Column (1) uses equation (2) but only including $V_{t-7}$ and $V_{t-14}$ . Column (2) in addition controls for log weekly deaths, log weekly cases and the linear time trend $T_t$ . Column (3) uses the first difference of log weekly first doses and includes deaths, cases and a time trend. We use a bandwidth of 50 days before and 35 days after the announcement date ( $T = 86$ ). <sup>†</sup>denotes that $H_0$ cannot be rejected at the 90% confidence level.

**Supplementary Table 9.** Time series estimates – alternative specifications

| p-values in [ ] | Outcome: log weekly vaccine first doses |  |  |  |
| --- | --- | --- | --- | --- |
| | deaths and hospitalizations | | binary $P_t$ | |
| | policy $\hat{\pi}$<br>(1) | trend change $\hat{\tau}_2$<br>(2) | policy $\hat{\pi}$<br>(3) | trend change $\hat{\tau}_2$<br>(4) |
| **Countries** |  |  |  |  |
| France | 0.468 ***<br>[0.000] | -0.009 **<br>[0.026] | 0.040<br>[0.321] | 0.004<br>[0.205] |
| Italy | 0.516 **<br>[0.035] | 0.014<br>[0.132] | 0.799 ***<br>[0.003] | -0.000<br>[0.967] |
| Germany | 0.385 ***<br>[0.000] | 0.007 ***<br>[0.000] | 0.274 ***<br>[0.001] | 0.007 ***<br>[0.002] |
| **Canadian provinces** |  |  |  |  |
| Quebec | 0.389 ***<br>[0.000] | -0.003<br>[0.387] | 0.016<br>[0.841] | 0.018 **<br>[0.027] |
| British Columbia | 0.626 ***<br>[0.000] | -0.007 ***<br>[0.000] | 0.566 ***<br>[0.000] | -0.003<br>[0.507] |
| Manitoba | 0.735 ***<br>[0.000] | 0.019 **<br>[0.018] | 0.586 ***<br>[0.002] | 0.012 *<br>[0.066] |
| Ontario | 0.322 ***<br>[0.003] | -0.002<br>[0.459] | 0.209 **<br>[0.022] | 0.002<br>[0.763] |
| Nova Scotia | 0.726 ***<br>[0.007] | 0.002<br>[0.782] | 0.470 ***<br>[0.003] | 0.026 ***<br>[0.003] |
| Alberta | 1.401 ***<br>[0.000] | -0.028 ***<br>[0.000] | 0.983 ***<br>[0.000] | -0.015 ***<br>[0.001] |
| New Brunswick | 1.220 ***<br>[0.000] | -0.014 **<br>[0.041] | 0.723 ***<br>[0.006] | -0.007<br>[0.431] |
| Saskatchewan | 0.809 ***<br>[0.000] | -0.009 *<br>[0.087] | 0.644 ***<br>[0.000] | -0.000<br>[0.945] |
| Newfoundland | 0.327<br>[0.146] | -0.001<br>[0.944] | 0.126<br>[0.534] | 0.011<br>[0.431] |
| Prince Edward Island | 0.323 *<br>[0.053] | -0.019 ***<br>[0.002] | 0.362 ***<br>[0.005] | -0.012 **<br>[0.047] |
| sample size for each row, T = 139 |  |  |  |  |

Notes: Time period – June 15 to October 31, 2021. All rows include 7-day and 14-day lags of the outcome variable. Columns (1) and (3) report the coefficient estimate $\hat{\pi}$ on the “mandate announced” policy variable $P_t$ in equation (2). Columns (2) and (4) report the estimate $\hat{\tau}_2$ on the post-announcement (interaction) time trend. Columns (1) and (2) use log weekly deaths and log average weekly hospitalizations as information; columns (3) and (4) use a binary (not weekly averaged) policy variable $P_t$ . P-values computed using Newey-West heteroskedasticity and autocorrelation robust (HAC) standard errors with 3 lags are in the square brackets. \*, \*\* and \*\*\* denote 10%, 5% and 1% significance (two-sided) respectively.

**Supplementary Table 10.** Counterfactuals – increase in first doses relative to no mandate as of Oct. 31, 2021

| | baseline:<br>cases and deaths | | deaths and<br>hospitalizations | binary $P_t$ |
| --- | --- | --- | --- | --- |
|  | total | 90% CI | total | total |
|  | (1) | (2) | (3) | (4) |
| **Countries (million doses)** |  |  |  |  |
| France | 4.59 | (2.47–6.25) | 4.80 | 2.86 |
| Italy | 6.48 | (2.67–8.14) | 6.23 | 5.78 |
| Germany | 3.47 | (3.06–3.81) | 3.52 | 3.37 |
| **Canadian provinces (thousand doses)** |  |  |  |  |
| Quebec | 235 | (111–298) | 177 | 229 |
| British Columbia | 158 | (106–191) | 113 | 157 |
| Manitoba | 32.8 | (24.2–39.1) | 35.9 | 32.9 |
| Ontario | 251 | (-41.6–386) | 192 | 210 |
| Nova Scotia | 37.4 | (32.2–41.2) | 34.7 | 37.4 |
| Alberta | 161 | (120–187) | 138 | 157 |
| New Brunswick | 35.4 | (27.5–40.9) | 33.7 | 26.7 |
| Saskatchewan | 49.7 | (40.3–56.2) | 41.8 | 54.6 |
| Newfoundland | 17.8 | (8.7–23.4) | 13.3 | 14.3 |
| Prince Edward Island | 1.3 | (-2.9–2.7) | -0.1 | 1.5 |
| Canada (total) | 979 | (425–1,266) | 779 | 920 |

Notes: The table displays the estimated increase in first-dose uptake, defined as the difference between total observed first-dose vaccinations and the mean estimated counterfactual first doses in the absence of vaccination mandate, as of October 31, 2021. Columns (1) and (2) use the baseline specification from Table 2 with log weekly cases and deaths as information. Column (3) uses log weekly deaths and log average weekly hospitalizations as information. Column (4) uses a binary policy variable instead of weekly average. The numbers for Canada in the last row are sums of the provincial gains.

**Supplementary Table 11.** Data sources

| Location | Vaccinations | Cases | Deaths |
| --- | --- | --- | --- |
| **Canadian provinces** |  |  |  |
| Alberta (AB) | link | link | link |
| British Columbia (BC) | link | link | link |
| Ontario (ON) | link | link | link |
| Quebec (QC) | link | link | link |
| Saskatchewan (SK) | link | link | link |
| Nova Scotia (NS) | link | link | link |
| Manitoba (MB) | link | link | link |
| New Brunswick (NB) | link | link | link |
| Newfoundland and Labrador (NL) | link | link | link |
| Prince Edward Island (PE) | link | link | link |
| **Countries** |  |  |  |
| France, Italy, Germany | link |  |  |

Notes: The data used in the paper are available at https://github.com/C19-SFU-Econ/dataV. The Canadian provincial data was last downloaded on Nov. 4, 2021. The country data was downloaded from OWID on Nov. 6, 2021. The BC and NS vaccinations data was manually collected from the official government bulletins. The NL vaccination data source is COVID-19CanadaOpenDataWorkingGroup, which archives official data from the provincial dashboards. In addition, the hospitalizations data used in the robustness analysis is from the COVID-19Tracker database (for the Canadian provinces, retrieved Nov. 12, 2021), OWID (for France and Italy, retrieved Nov. 6, 2021), and the RobertKochInstitute (for Germany, retrieved Dec. 14, 2021).

**Supplementary Table 12.** Mandates scope

| Canadian Provinces |  |
| --- | --- |
| Quebec | Health and social facilities (hospitals, rehabilitation, etc.); Private seniors' residences; indoor sports and physical activities; outdoor organized sports and physical activities involving frequent or prolonged contact; Outdoor events and festivals with more than 50 spectators or participants; Sporting events or shows (play, concert, musical or comedy shows etc.) in outdoor stadiums or outdoor stage; Agricultural fairs; Festivals or celebrations; Walks, marathons, cycling circuits; Golf tournaments; Immersive or walking tours; Performance venues; Stadiums and arenas; Auditoriums; Movie theatres; Indoor sport events at which the number of spectators exceeds 250 people seated in assigned seats; Bars and restaurants, including patios; Fast food restaurant dining rooms, including patios; Nightclubs; Microbreweries; Distilleries; Shopping mall food courts and food stores; Arcades, theme parks, amusement parks and centres, recreation centres, and water parks; Casinos and gambling halls, including bingo; Tourist and recreational cruises; Conventions and conferences. |
| British Columbia | Indoor ticketed sporting events with more than 50 people; Indoor concerts, theatre, dance and symphony events with more than 50 people; Licensed restaurants and cafes and restaurants and cafes that offer table service (indoor and patio dining); Liquor tasting rooms in wineries, breweries or distilleries; Pubs, bars and lounges (indoor and patio); Nightclubs, casinos and movie theatres; Gyms, exercise and dance facilities or studios; Businesses offering indoor exercise/fitness; Indoor adult group and team sports for people 22 or older; Indoor organized events with more than 50 people (e.g., weddings, funeral receptions, organized parties, conferences, trade fairs and workshops); Indoor organized group recreational classes and activities with more than 50 people; Post-secondary student housing; Spectators at indoor youth sporting events with more than 50 people. |
| Manitoba | Indoor and outdoor ticketed sporting events and concerts; Indoor theatre/dance/symphony events; Restaurants (indoor and patio dining); Nightclubs and all other licensed premises; Casinos, bingo halls and VLT lounges; Movie theatres; Fitness centres, gyms and indoor sporting and recreational facilities (excluding youth recreational sport); Organized indoor group recreational classes and activities and indoor recreational business. |
| Ontario | Restaurants, bars and other food and drink establishments; food or drink establishments with dance facilities, including nightclubs, restoclubs, and outdoor areas of these establishments; meeting and event spaces; facilities used for sports and recreational fitness activities and personal physical fitness training, including: gyms, fitness, sporting and recreational facilities; pools; leagues, sporting events; waterparks; indoor areas of facilities where spectators watch events; casinos, bingo halls and other gaming establishments; concert venues, theatres and cinemas; bath-houses, sex clubs and strip clubs; horse racing tracks, car racing tracks and other similar venues; commercial film and TV productions where there is a studio audiences; any of the following outdoor areas that have a normal capacity of 20,000 or more people: outdoor meeting and event spaces; outdoor facilities used for sports and recreational fitness activities; outdoor concert venues, theatres and cinemas. |
| Alberta<sup>1</sup> | Restaurants and food courts; Nightclubs, casinos, bingo halls, VLT lounges; Entertainment and recreation centres (e.g., bowling, racing, arcades, billiards halls, etc.); Museums, art galleries; Movie theatres; Recreation facilities for physical, performance or recreational activity; Conferences, meeting spaces, halls, and rented space; Weddings and funerals held in public facilities; Professional sporting or performance events (spectator); Indoor adult sport and performance activities (participants); Private social events held in public facilities; Adult recreational activities (e.g., classes, groups); Amenities in hotels and condos, including fitness rooms, pools and game rooms. |
| Nova Scotia | Full-service restaurants where patrons sit at tables to be served, both indoors and on patios; food establishments (like fast food and coffee shops) where people sit to eat and drink, both indoors and on patios (not including food courts, takeout, drive-thru or delivery); liquor licensed (drinking) establishments (like bars, wineries, distillery tasting rooms, craft taprooms and liquor manufacturers), both indoors and on patios; casinos and gaming establishments, both indoors and on patios; fitness establishments (like gyms and yoga studios) and sport and recreation facilities (like arenas, pools and large multipurpose recreation facilities); businesses and organizations offering indoor and outdoor recreation and leisure activities (like climbing facilities, dance classes, escape rooms, go-carts, indoor arcades, indoor play spaces, music lessons, pottery painting, shooting ranges and outdoor adventure); indoor and outdoor festivals, special events and arts and culture events (like theatre performances, concerts and movie theatres); indoor and outdoor sports practices, games, competitions and tournaments (participants and spectators); indoor and outdoor extracurricular school-based activities, including sportsbus, boat and walking tours; museums, Art Gallery of Nova Scotia and public library programs; indoor and outdoor events and activities like receptions, social events, conferences and training that are hosted by a business or organization; indoor and outdoor wedding ceremonies and funerals (including receptions and visitation) that are hosted by a business or organization; community meetings in rental spaces or where the public may be present; training hosted by a business or organization. |
| New Brunswick | Indoor festivals, performing arts and sporting events; Indoor and outdoor dining and drinking at restaurants, pubs and bars; Nightclubs, amusement centres, pool halls, bowling alleys and casinos; Movie theatres; Gyms, indoor group exercise, indoor pools and recreation facilities; Indoor organized gatherings like weddings, funerals, parties, conferences and workshops; Indoor organized group recreational classes and activities; Visiting a long-term care facility. |
| Saskatchewan<sup>2</sup> | Restaurants, including in hotels or other lodgings (incl. outdoor patios); Nightclubs, bars, taverns buses and other establishments that serve alcohol; theatres; cinemas; bingo halls, casinos and other gaming establishments; concerts; live-music venues; fitness centres and gyms; stand-alone liquor and cannabis retail sales locations; and facilities hosting sporting events where tickets are required that have GST charged. |
| Newfoundland and Labrador | Gatherings of any size held for socializing, celebration, ceremony or entertainment hosted at a recognized business or organization, a rental room, community centre, or other venues used to host gatherings (e.g. weddings, funerals, birthday parties, baby showers, faith-based gatherings); Conferences, conventions and trade shows; Arenas; Indoor gyms and fitness facilities, yoga studios, and dance studios; Places where sports or recreational activities are practiced indoors; Places where group music, art, dance, and drama activities are practiced indoors, including bands, choirs, dance and music classes; Indoor entertainment facilities (arcades, trampoline parks, bowling alleys, billiard halls, golf, laser tag, indoor playgrounds, and paintball); Bars and lounges; Restaurants (indoor seated dining only, including food courts; does not apply to outdoor patios, take-out, delivery, or drive-thru services); Cinemas and performance spaces; Bingo halls; Personal service establishments including spas, esthetic services, hair salons, barber shops, body piercing, tattooing and tanning salons; Long-term care homes, personal care homes, assisted living facilities, community care homes (visitors only). |
| Prince Edward Island | Indoor and outdoor organized gatherings/events, including sporting events and recreational activities, and spectators at youth sporting events or recreational activities; Concerts, and arts, theatre, and music events; Wedding and funeral receptions and wakes; Conferences, trade fairs and workshops; Group activities and classes like pottery, art and choir; Adult group and team sports for people 19 years of age and older; Food premises and licensed premises (indoor and patio dining), including: Restaurants, Cafes, Bars, Liquor tasting rooms in wineries, breweries or distilleries; Casinos and movie theatres; Indoor gyms, exercise/dance facilities/studios, swimming pools and skating rinks; Outdoor facilities for organized gatherings/events; Arcades and bowling alleys; Meetings where members of the general public may be present. |

| Countries |  |
| --- | --- |
| France<sup>3</sup> | Marquees, theaters, sporting or cultural performance venues, conference rooms; trade shows and exhibition fairs; outdoor establishments including zoological, amusement and theme parks; stadiums, sports establishments, swimming pools, sports halls; casinos, gaming halls and "bowling alleys"; outdoor sit/stand festivals; cinemas and theaters; monuments, museums and exhibition halls; libraries (excluding university and specialized libraries); sports competition; other events, cultural, sporting, fun or festive, organized in public space or in a place open to the public; worship establishments for events not of a religious nature; ships and boats, cruise ship type; discotheques, dance clubs and bars; fairgrounds, from a threshold of 30 stands or attractions; commercial catering activities (bars and restaurants, including on terraces); trade fairs and exhibitions, and professional seminars; health, social and medico-social services and establishments; long-distance travel by inter-regional public transport (domestic flights, TGV journeys, Intercity and night trains, inter-regional coaches); department stores and shopping centers of more than 20,000 m<sup>2</sup>. |
| Italy<sup>3</sup> | Travel between different regions by air, train, ship, ferry or coach; Restaurants, bars, ice cream parlours and pastry shops for consumption at table indoors; Performances open to the public, sporting events, both outdoors and indoors; Museums and places of culture, shows; Swimming pools and gyms; Private parties, such as wedding receptions; Festivals and trade fairs; Conventions and congresses; Spas and fitness centres; Gaming halls and betting shops, bingo halls and casinos. |
| Germany<sup>3</sup> | Visitor access to hospitals, nursing and care homes and assisted living facilities for the disabled; Access to indoor cafés and restaurants; Participation in indoor events and celebrations (e.g. information events, cultural events or sporting events); Using personal care services (e.g. hairdressers, beauticians, cosmetic services); Indoor sports (e.g. at gyms, swimming pools or sports centres); Accommodation (hotels, guesthouses): a test is to be taken for arrival and twice a week for the duration of the stay. |

Notes: During the studied period: 1. Alberta's mandate allows a recent negative COVID-19 test in lieu of vaccination and allows businesses to opt out of the proof of vaccination requirement and instead be subject to stricter COVID-19 prevention protocols, e.g., capacity limits, distancing, etc. 2. Saskatchewan's mandate allows a recent negative COVID-19 test in lieu of vaccination. 3. France's, Italy's and Germany's mandates allow a recent negative COVID-19 test or a past positive test in lieu of vaccination.

